# A deep learning ECG model for localization of occlusion myocardial infarction

**DOI:** 10.1101/2025.09.11.25335407

**Authors:** Stefan Gustafsson, Antônio H. Ribeiro, Daniel Gedon, Petrus E. O. G. B. Abreu, Nicolas Pielawski, Gabriela M. M. Paixão, Antonio Luiz P. Ribeiro, Daniel Lindholm, Thomas B. Schön, Johan Sundström

## Abstract

Rapid identification and localization of an acute coronary occlusion are vital to prevent myocardial damage, yet reliance on ST-segment ECG criteria misses many acute occlusion myocardial infarctions (OMI) and triggers unnecessary acute angiographies. Here, we trained and validated a deep learning model using 540,372 emergency ECGs paired with definitive catheterization outcomes. The model achieved a C-statistic of >0.95 for OMI and >0.87 for non-OMI infarctions and could localize culprit lesions in the left main/LAD, LCX, and RCA coronary branches, which can guide the angiographer. Performance was similar across age, sex, and ECG hardware subgroups. Obviating dependence on ST-elevations and troponins, this model for identification and localization of OMI has the potential to shorten time to reperfusion of an acute coronary occlusion and save resources. Because human oversight of OMI detection on the ECG is limited, randomized clinical trials with patient-relevant outcomes are warranted.

---

Acute myocardial infarction is the leading cause of death globally.^1^ When a patient presents with a suspected acute coronary syndrome, the goal is to decide if the patient needs to be urgently transported to a coronary intervention lab or if the angiography can wait, e.g. until office hours. If the coronary artery is acutely occluded, time equals dying myocardium. The electrocardiogram (ECG) is, therefore, of great utility because it can often be obtained already in the ambulance and could herald an acute occlusion before permanent myocardial damage entails. The current ECG paradigm mainly utilizes the ST-segment for these decisions, but ST-elevations are only present in a fraction of acute occlusion myocardial infarctions (OMI),^2^ ST-elevations can have other causes than OMI, and physicians disagree in their interpretations of ST-elevations.^3,4^ This leads to lost time,^5,6^ lost lives,^2^ high costs and risks associated with unnecessary coronary angiographies. Recent dramatic advances in ECG interpretation using machine learning offer the potential to better identify an OMI, potentially saving time, lives and costs.^7^ Machine learning models using parameterized^8^ or raw^9^ ECG data have shown promise for OMI classification. The ECG can also be useful for determining the localization of pathologies, and machine learning models have been shown to have the capacity to localize a non-occlusive coronary stenosis, i.e., the potential to localize coronary artery narrowing in the absence of a complete occlusion.^10^ Some patients have chronic coronary occlusions in addition to an ongoing acute OMI, making the choice of which coronary occlusion to intervene on difficult but critical.^11^ Correctly identifying the localization of the acute occlusion would therefore be of great clinical value. We developed and validated such a machine learning model for the identification of OMI and its localization, by using more than half a million ECGs and detailed anatomical coronary angiography and intervention results in emergency department patients.

We utilized 540,372 ECGs from 465,471 emergency department (ED) visits and 225,824 patients in the Swedish Emergency Department Database (SwED; Supplementary Figure 1).^12^ Of these ECGs, 1,583 (0.3%), 4,279 (0.8%), and 534,510 (98.9%) cases were labeled at time of coronary catheterization by the coronary angiographer as OMI, nOMI (myocardial infarction not classified as OMI), and control (no myocardial infarction), respectively. OMI was defined as the presence of a newly formed coronary total occlusion or very close to total occlusion (TIMI blood flow 0). Of those with an OMI, 595 (37.6%), 359 (22.7%), and 629 (39.7%) had the left main or left anterior descending artery (LM/LAD), the left circumflex artery (LCX), and the right coronary artery (RCA) as the culprit vessel (the acutely blocked artery responsible for this myocardial infarction), respectively (Figure 1, panels A and B). We used 70% of the SwED data (data split on patients, not ECGs) for training, 10% for validation, and 20% for testing (one random test set, and one temporal test set with patients whose first visit occurred in the final year of the study period). The numbers in each outcome class (defined in Supplementary Figure 2) and data split are shown in Supplementary Table 1, and the numbers in each subclass over time are shown in Supplementary Figures 3 and 4. The number of samples in each sub class increased over the years, likely due to multiple factors, e.g. including expanded ECG data coverage and a growing proportion of NSTEMI patients managed invasively (with available angiography). The clinical characteristics by OMI class are shown in Table 1 for all data splits of SwED combined, showing that the OMI cases were on average younger than the nOMI cases, had a higher proportion of men, and had less known underlying cardiovascular comorbidities and use of cardiovascular drugs at the time of the ED visit. The OMI cases also had markedly elevated levels of cardiac troponin levels, an elevated risk of death within 30 days (Table 1), and all underwent percutaneous coronary intervention (PCI) within the first week after ED admission (Supplementary Figure 5).

**Table 1.**
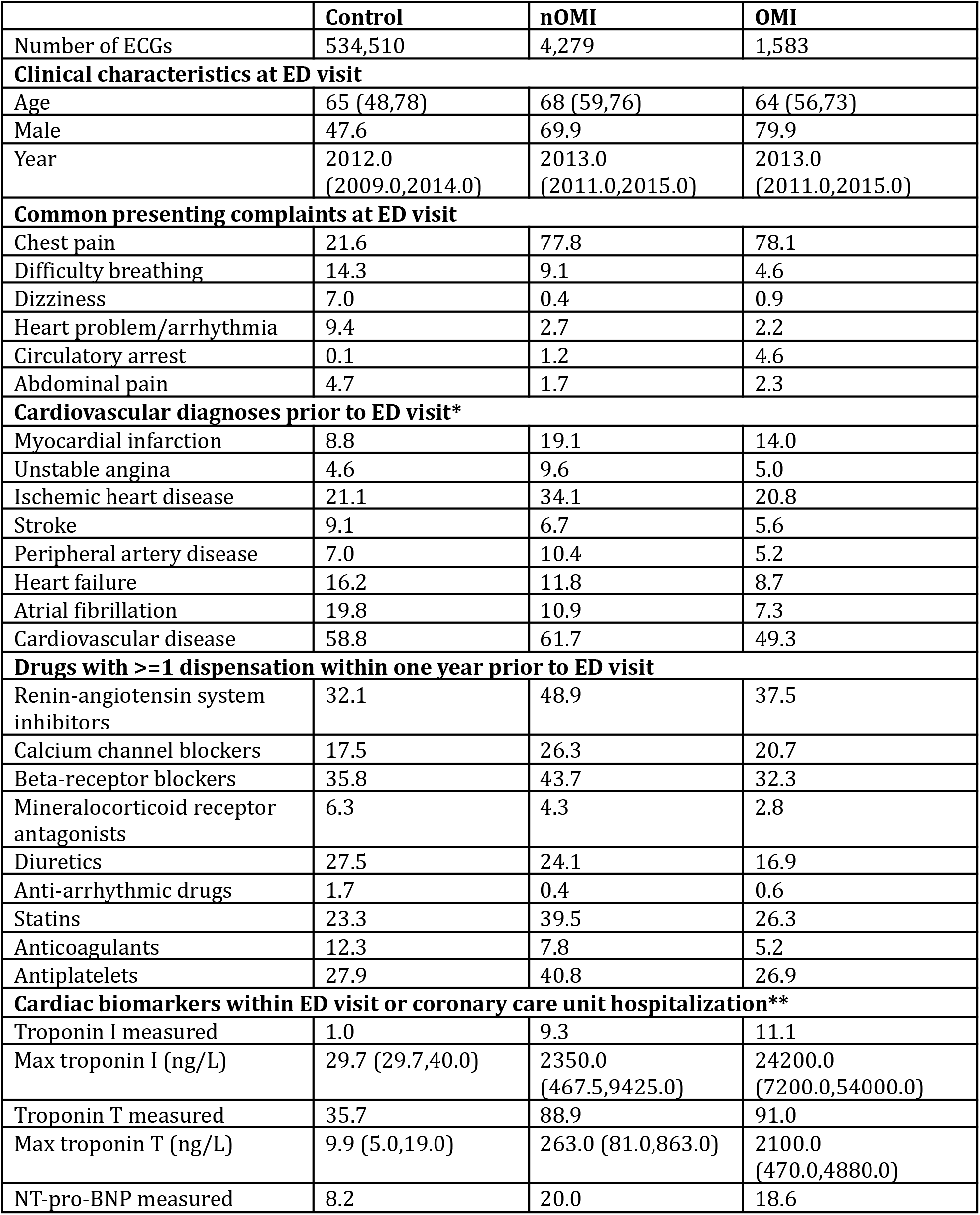
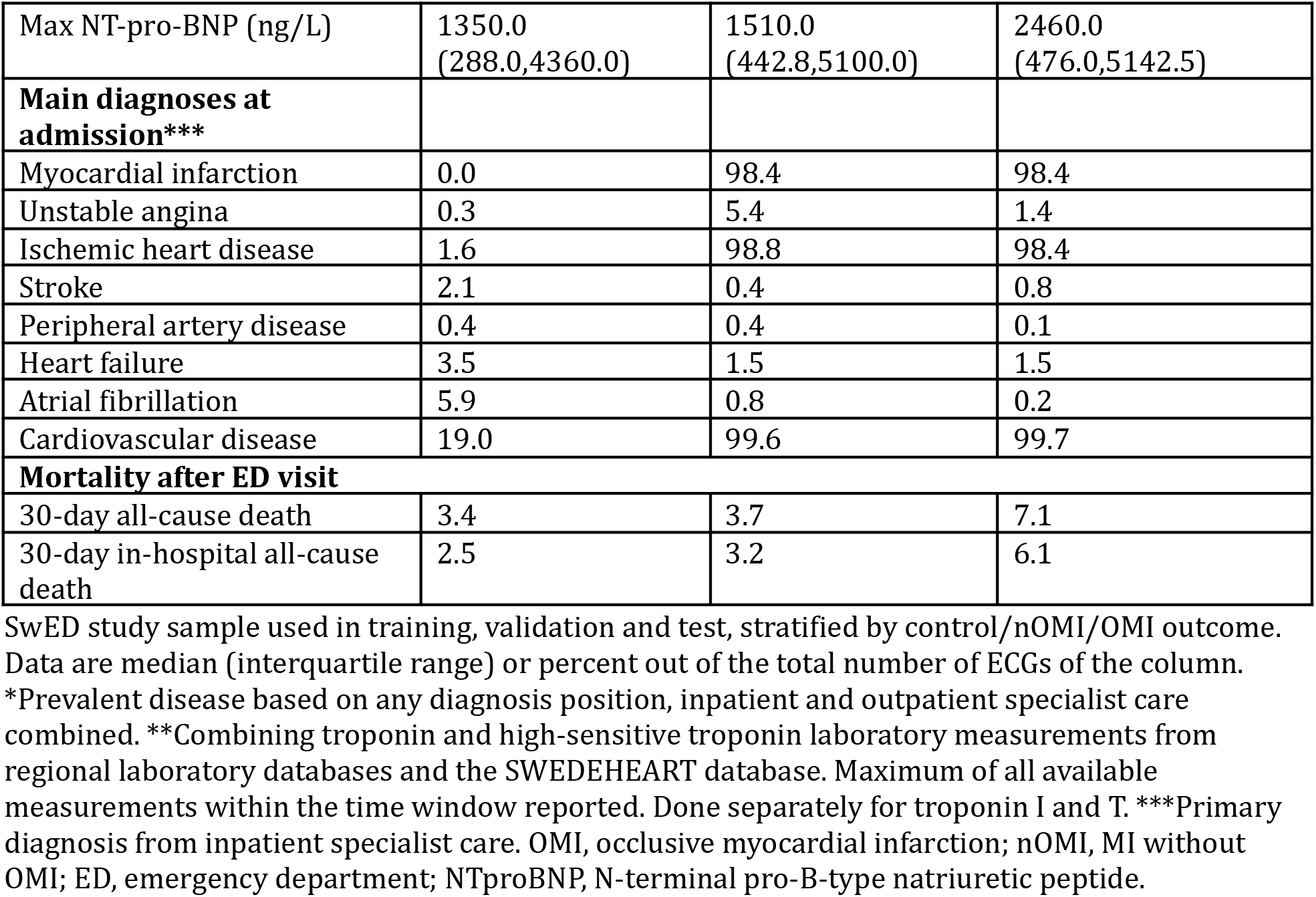
Clinical characteristics of the study sample

|  | Control | nOMI | OMI |
| --- | --- | --- | --- |
| Number of ECGs | 534,510 | 4,279 | 1,583 |
| **Clinical characteristics at ED visit** |  |  |  |
| Age | 65 (48,78) | 68 (59,76) | 64 (56,73) |
| Male | 47.6 | 69.9 | 79.9 |
| Year | 2012.0<br>(2009.0,2014.0) | 2013.0<br>(2011.0,2015.0) | 2013.0<br>(2011.0,2015.0) |
| **Common presenting complaints at ED visit** |  |  |  |
| Chest pain | 21.6 | 77.8 | 78.1 |
| Difficulty breathing | 14.3 | 9.1 | 4.6 |
| Dizziness | 7.0 | 0.4 | 0.9 |
| Heart problem/arrhythmia | 9.4 | 2.7 | 2.2 |
| Circulatory arrest | 0.1 | 1.2 | 4.6 |
| Abdominal pain | 4.7 | 1.7 | 2.3 |
| **Cardiovascular diagnoses prior to ED visit\*** |  |  |  |
| Myocardial infarction | 8.8 | 19.1 | 14.0 |
| Unstable angina | 4.6 | 9.6 | 5.0 |
| Ischemic heart disease | 21.1 | 34.1 | 20.8 |
| Stroke | 9.1 | 6.7 | 5.6 |
| Peripheral artery disease | 7.0 | 10.4 | 5.2 |
| Heart failure | 16.2 | 11.8 | 8.7 |
| Atrial fibrillation | 19.8 | 10.9 | 7.3 |
| Cardiovascular disease | 58.8 | 61.7 | 49.3 |
| **Drugs with $\geq 1$ dispensation within one year prior to ED visit** |  |  |  |
| Renin-angiotensin system inhibitors | 32.1 | 48.9 | 37.5 |
| Calcium channel blockers | 17.5 | 26.3 | 20.7 |
| Beta-receptor blockers | 35.8 | 43.7 | 32.3 |
| Mineralocorticoid receptor antagonists | 6.3 | 4.3 | 2.8 |
| Diuretics | 27.5 | 24.1 | 16.9 |
| Anti-arrhythmic drugs | 1.7 | 0.4 | 0.6 |
| Statins | 23.3 | 39.5 | 26.3 |
| Anticoagulants | 12.3 | 7.8 | 5.2 |
| Antiplatelets | 27.9 | 40.8 | 26.9 |
| **Cardiac biomarkers within ED visit or coronary care unit hospitalization\*\*** |  |  |  |
| Troponin I measured | 1.0 | 9.3 | 11.1 |
| Max troponin I (ng/L) | 29.7 (29.7,40.0) | 2350.0<br>(467.5,9425.0) | 24200.0<br>(7200.0,54000.0) |
| Troponin T measured | 35.7 | 88.9 | 91.0 |
| Max troponin T (ng/L) | 9.9 (5.0,19.0) | 263.0 (81.0,863.0) | 2100.0<br>(470.0,4880.0) |
| NT-pro-BNP measured | 8.2 | 20.0 | 18.6 |
| Max NT-pro-BNP (ng/L) | 1350.0<br>(288.0,4360.0) | 1510.0<br>(442.8,5100.0) | 2460.0<br>(476.0,5142.5) |
| **Main diagnoses at admission\*\*\*** |  |  |  |
| Myocardial infarction | 0.0 | 98.4 | 98.4 |
| Unstable angina | 0.3 | 5.4 | 1.4 |
| Ischemic heart disease | 1.6 | 98.8 | 98.4 |
| Stroke | 2.1 | 0.4 | 0.8 |
| Peripheral artery disease | 0.4 | 0.4 | 0.1 |
| Heart failure | 3.5 | 1.5 | 1.5 |
| Atrial fibrillation | 5.9 | 0.8 | 0.2 |
| Cardiovascular disease | 19.0 | 99.6 | 99.7 |
| **Mortality after ED visit** |  |  |  |
| 30-day all-cause death | 3.4 | 3.7 | 7.1 |
| 30-day in-hospital all-cause death | 2.5 | 3.2 | 6.1 |

SwED study sample used in training, validation and test, stratified by control/nOMI/OMI outcome. Data are median (interquartile range) or percent out of the total number of ECGs of the column.
\*Prevalent disease based on any diagnosis position, inpatient and outpatient specialist care combined. \*\*Combining troponin and high-sensitive troponin laboratory measurements from regional laboratory databases and the SWEDEHEART database. Maximum of all available measurements within the time window reported. Done separately for troponin I and T. \*\*\*Primary diagnosis from inpatient specialist care. OMI, occlusive myocardial infarction; nOMI, MI without OMI; ED, emergency department; NTproBNP, N-terminal pro-B-type natriuretic peptide.

**Figure 1.**
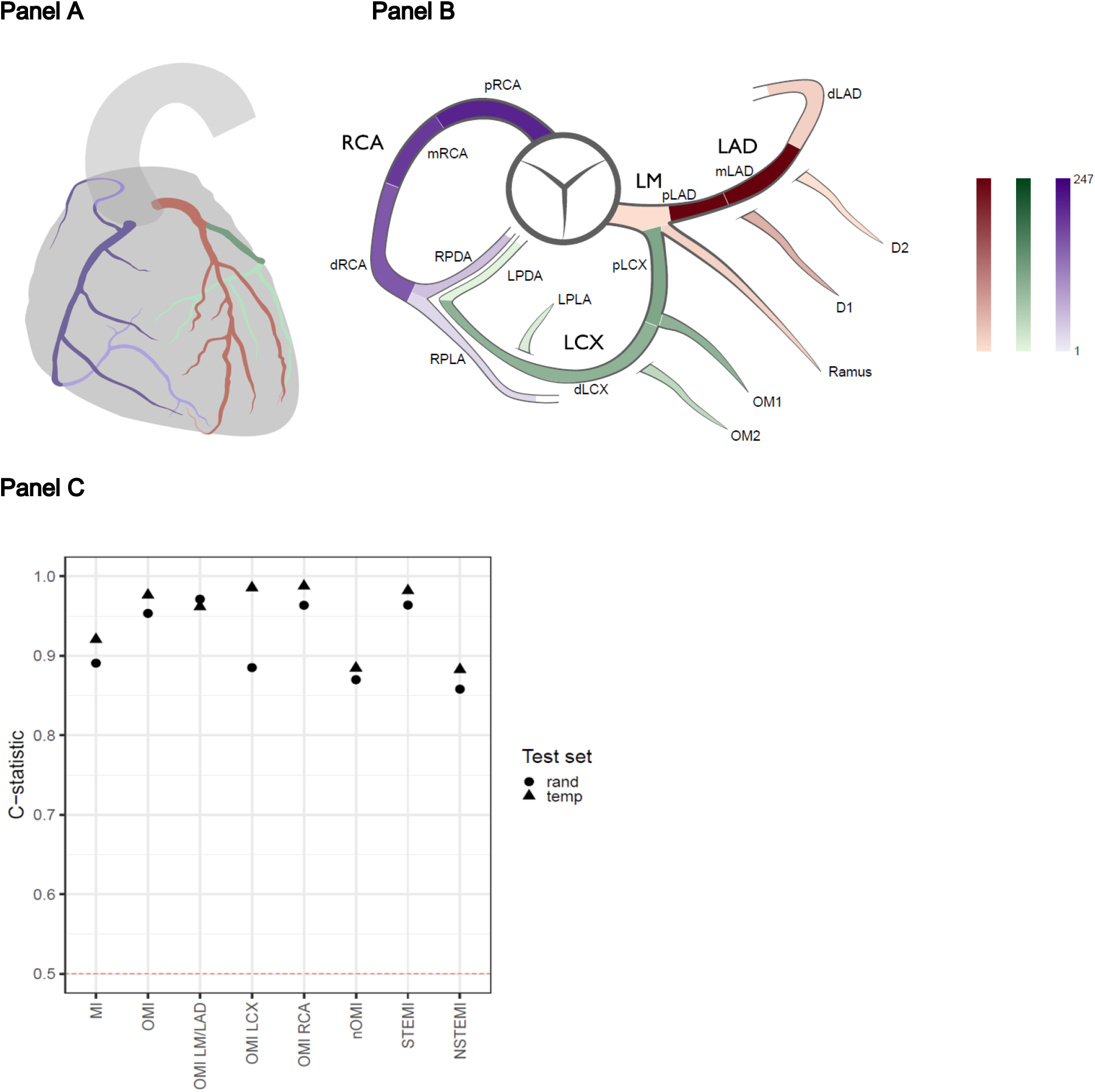
Localization of occlusion myocardial infarctions and model performance. **Panel A** shows an anatomical heart with the three main branches of the coronary tree, where the model tries to predict the location of an occlusion. RCA (right coronary artery) in purple, LM/LAD (left main coronary artery+left anterior descending artery) in red, and LCX (left circumflex artery) in green, where a lighter color corresponds to a coronary artery that is hidden behind the current view of the heart. **Panel B** shows a schematic illustration of the coronary artery segments centered around the aorta. The same color scheme is used as in Panel A, but the color intensity corresponds to how frequent occlusive myocardial infarctions were in the given segment. **Panel C** shows the discriminative performance (C-statistic) when comparing a given class (x-axis) with all other classes in the random or temporal test set. The dashed horizontal line represents a random guess.

The model performance in the random and temporal test sets is presented in Figure 1, panel C (discriminatory performance of the super-classes of the model), Supplementary Figure 6 (discriminatory performance of the sub-classes of the model), and Supplementary Table 2 (metrics of both discrimination and calibration for all super- and sub-classes, including model uncertainty over ten different initialization seeds). Supplementary Figure 7 presents the receiver operating characteristic (ROC) and precision-recall curves across both test datasets. These above-mentioned results present metrics comparing one given class vs all the other classes (OvA) or multiclass comparisons. Supplementary Figure 8 presents metrics for one vs one (OvO) comparisons. In the SwED test sets, the model demonstrated excellent discriminatory performance for the OMI superclass, achieving an OvA C-statistic of >0.95. For the nOMI superclass, the model yielded a OvA C-statistic of >0.87. The model also performed well in identifying the OMI culprit vessel, but with variability in the results for some specific subclasses and test splits. This applied to OvA comparisons; the discriminatory performance was poor for one OvO comparison, OMI LCX vs OMI RCA. We also tested model performance in the publicly available PTB-XL test dataset (European) and the CODE-II test dataset (Brazilian); the model achieved excellent performance (Supplementary Table 3) in differentiating STEMI vs non-infarctions and NSTEMI (the only available labels in these test sets). Calibration plots indicated acceptable agreement between predicted probabilities and observed outcomes, as shown in Supplementary Figure 9. The resolution of the binned calibration plots was sometimes low due to few cases in the outcome classes.

To assess the robustness of our model and rule out the influence of proxy variables, we stratified performance by several potential confounders. Stratified C-statistics are presented in Supplementary Figure 10 for the following subsets: age tertiles, sex, did the patient visit the emergency department at Karolinska Hospital (main source of data) or another emergency department in the Stockholm region, ECGs recorded using the most common machine type (MAC55) or not, ECGs recorded using the most common software (v237) or not, prior hospitalisation due to cardiovascular disease, and prevalent left bundle-branch block (LBBB). The discriminatory performance was mostly comparable across strata, but some differences should be highlighted. The model performed better in patients free from LBBB, younger patients, and patients free from prior cardiovascular disease hospitalisations.

Some comorbidities were over-or underrepresented among certain high probability misclassifications as shown in Supplementary Table 4. A manual review (GMMP, consultant cardiologist) of CODE-II ECGs misclassified with a high probability suggested that many discrepancies were due to incorrect reference diagnoses. In several cases, the reviewer agreed more with the model than with the original label. Among the false positives, all ECGs showed ST-segment elevation, with eight displaying classic STEMI patterns and two showing features of pericarditis or a dyskinetic segment. Half of the false negatives showed clear STEMI patterns, while the rest included confounding features such as LBBB, subendocardial ischemia, left ventricular hypertrophy, premature ventricular contractions, sinus tachycardia, and atrial fibrillation with high ventricular rate, which may explain the model’s low confidence (Supplementary Table 5).

The model displayed excellent discrimination but lower average precision; it seemed unsure on the right occasions, e.g. with challenging ECGs such as with technical noise that the normalization had not removed, in cases with very early ECGs on arrival at the ED or very late when thrombolysis treatment or even PCI hay have been performed, and with possible (but not probable) STEMI in PTB-XL. A manual review (JS, consultant cardiologist) of test set patients with multiple ECGs at the same visit but where the model outputs very different probabilities showed that the ECGs given a high OMI probability indeed had a clear infarction pattern whereas ECGs with a low probability often had issues such as a much weaker infarction pattern or major technical noise.

Weaknesses include test sets’ geographical similarity to the training set with all ECGs in a similar format albeit with some differences in machines, analysis software version, and sampling frequency; no availability of independent test sets with same OMI labels; test sets from other geographical regions (likely with different machine setup) only include STEMI labels. Certain subgroups remain an issue; perimyocarditis still^12^ raises some confusion for the model, and discrimination of nOMI in the LBBB subgroup is poor but better than chance.

Strengths of this study include a strict OMI definition based solely on catheterization data that can differentiate acute from chronic coronary occlusions. OMI definitions that rely on cardiac troponin levels are suboptimal; STEMI cases are rushed past the ED to the catheterization lab without stopping for cardiac troponin testing; troponin levels are low in the most acute phase; and the acuity of less than total or near-total occlusions is unknown. Our model does not rely on troponins or any other external dependencies that may lead to delays. Most myocardial infarction deaths occur in low- and middle-income countries, where timely access to reperfusion therapy is often limited. In these settings, accurate and affordable tools such as machine learning-based ECG interpretation could play a vital role in increasing access to early diagnosis and life-saving treatment^13^. An unanticipated observation is that our model’s STEMI predictions^12^ outperform a STEMI model trained on the CODE-II dataset itself, when testing it on the CODE-II dataset. Hence, a model trained on human labels (the CODE-II data) can reach the best conceivable human level, while a model trained on objective outcome labels (our coronary catheterization data) can acquire beyond-human capabilities, even when the test set is based on human labels.

We herein developed and validated a first machine learning model for identification and localization of OMI. The clinical usefulness of such a model can be substantial as it can (1) identify OMI patients that are in need of urgent revascularization, and avoid acute activation of the catheterization lab if there is no OMI; and (2) guide the angiographer to the critical coronary branch, in the case of concurrent chronic occlusions^11^. This is a major development over the current STEMI/NSTEMI paradigm, as ST-elevations are only present in a fraction of OMIs in need of acute revascularization^2^, leading to unnecessary delays^5,6^, deaths^2^, and costs. Prior machine learning models using ECG data^8,9^ have shown high performance for classification of OMI. Our model reached similar or higher performance, and can additionally point out the localization of the occlusion.

These findings further strengthen the notion that OMI is a potentially useful clinical entity, and the potential usefulness of machine learning models for its timely detection and localization. Because human oversight of OMI detection on the ECG is limited, randomized clinical trials with patient-relevant outcomes are needed before clinical deployment of such models.

## Supporting information

All supplementary materials

## Inclusion and Ethics

The inclusion criteria were kept as wide as possible: only minors and patients without a personal identification number were excluded. The study was approved by the Swedish Ethics Authority, accession numbers 2022-07108-01, 2023-05042-02, 2023-09-22, and 2024-04058-02.

## Data availability statement

The data that support the findings of this study are available from the Swedish Board of Health and Welfare and the included healthcare regions, but restrictions apply to the availability of these data, which were used under license for the current study.

## Code availability statement

All code together with the parameter/hyperparameter estimates of the presented models is available upon request.

## Online methods

### Main study sample

The main study sample used in training and validation of the model was obtained from a subset of the Swedish Emergency Department Database (SwED), as described in a previous study of the prediction of ST-elevation and non-ST-elevation myocardial infarction (STEMI and NSTEMI).^12^ In brief, the SwED sample includes patients >= 18 years old with available ED data from multiple emergency departments in Sweden, between 2003 and 2017. The sample was linked to national registries (the patient [inpatient and specialized outpatient], prescribed drug, and death registries), national quality registries (SWEDEHEART [Swedish Web-system for Enhancement and Development of Evidence-based care in Heart disease Evaluated According to Recommended Therapies; a Swedish nation-wide quality register] sub-registries RIKS-HIA [Register of Information and Knowledge About Swedish Heart Intensive Care Admissions] and SCAAR [Swedish Coronary Angiography and Angioplasty Registry]), as well as a regional database of ECGs (Karolinska ECG database) and electronic health records.

ECG availability restricted the present study to the Karolinska ECG database, Stockholm, Sweden and the years covered by all required data sources at time of the SwED data extraction was limited to 2005-2016. The Karolinska ECG database mainly consists of patients admitted to the Karolinska Hospital, Stockholm, Sweden (85%) but also includes other Region Stockholm hospitals such as Södertälje and Norrtälje. Hence, the study sample included all adult patients who presented to the emergency department (ED) in Region Stockholm, Sweden, between 2005 and 2016, where a standard 12-lead ECG was recorded within 24 hours of their visit. The inclusion and exclusion criteria used to define the study sample are described in Supplementary Figure 1, which were largely consistent with the prior study^12^. Notable differences were the requirement that all myocardial infarction (MI) cases have undergone angiography within seven days of their ED visit (in order to differentiate between acute and chronic occlusions and to determine localization), the additional inclusion of patients with prevalent left bundle branch block (LBBB; as this is a non-negligible fraction of all acute occlusions that any MI model should be able to identify), and the inclusion of controls from the years 2005-2006 (the granular MI subclasses described below were not recorded for these years, hence only controls were included).

Ethical approval was obtained from the Swedish Ethical Review Agency (application number 2020-01654), and informed consent was waived as per applicable ethical guidelines.

### Exposures and outcomes

As in the previous study^12^, high-quality data on exposures and outcomes were available from emergency department discharge records, electronic health records, the SWEDEHEART registry, and linked hospitalizations, with patient records joined using the Swedish personal identifier number.

The exposures included digital ECG data, age at time of the ED visit, and sex. The ECG recordings were available in the GE MUSE (v9) XML format, with each lead represented by a numerical signal vector together with detailed meta-data of the recording. Standard 10-second, 12-lead ECG recordings sampled at 250 to 500 Hz were used; eight leads were retained (I, II, V1-V6) as four of the standard leads are linear combinations of these and thus redundant. All ECGs were resampled to 400 Hz and normalized as described previously^12^.

The outcome labels are primarily defined using data from the SWEDEHEART quality registry where an overall data accuracy of around 96% has been reported from monitor visits. Further, SWEDEHEART has a coverage of 100% for Swedish patients undergoing angiography, which is an inclusion criteria for the myocardial infarction cases in the present study.^14^ The study extended the three outcome classes of the prior study^12^ (control, STEMI, and NSTEMI) into ten mutually exclusive MI subclasses, incorporating additional granularity regarding OMI and culprit vessel localization (definitions described below). Additionally, controls were stratified into those with and without perimyocarditis, a subgroup previously found to be frequently misclassified as STEMI.^12^ The MI subclasses included:

1. Control with perimyocarditis
2. Control without perimyocarditis
3. NSTEMI, nOMI
4. NSTEMI, OMI, LM/LAD
5. NSTEMI, OMI, LCX
6. NSTEMI, OMI, RCA
7. STEMI, nOMI
8. STEMI, OMI, LM/LAD
9. STEMI, OMI, LCX
10. STEMI, OMI, RCA

OMI was defined as a newly formed occlusion with complete or close to complete blockage requiring more urgent intervention as opposed to old chronic occlusions that have slowly been built up (where the heart, to some extent, might have adapted, by forming collateral vessels bypassing the blockage). OMI was defined based on the SWEDEHEART SCAAR variable OCKL. The culprit vessel of the OMI was defined as the segment marked as having the newly formed occlusion. The SCAAR database contains information on the coronary segments affected, with twenty segment labels according to a modified version of a previous definition^15^. The fraction of MI cases with an occlusion in some of these segments is very small, and it is sufficient to guide the angiographer to the correct main branch; therefore the segments were grouped into the three main branches: left main coronary artery together with the left anterior descending artery and the intermediary (LM/LAD), left circumflex artery (LCX), and right coronary artery (RCA) as illustrated in Figure 1, which shows all segments together with the main branches.

Instead of excluding patients with prevalent LBBB as in the STEMI/NSTEMI study, the presence of LBBB was incorporated as a separate outcome label that could co-occur with the ten MI classes. LBBB classification was based on the LBBB predictions from an external, machine learning model trained on Brazilian data, which has been shown to outperform cardiology resident medical doctors in the classification of LBBB from ECGs.^16^ The model predictions were supplemented with prevalent ICD10:I44.6-I44.7 diagnoses from the patient registry and SWEDEHEART annotations of LBBB. A patient was classified as having an LBBB at the ED visit if all ECGs of the ED visit had a Pr(LBBB)>0.5 (cutoff selected by visual inspection of Supplementary Figure 11) or if a prevalent LBBB diagnosis was present in any of the other data sources.

### Outcome combinations

For the ten non-overlapping MI outcome sub classes, several super categories were of particular interest:

- OMI = combination of all OMI classes
- OMI LM/LAD = combination of all OMI classes with LM/LAD as the culprit vessel
- OMI LCX = combination of all OMI classes with LCX as the culprit vessel
- OMI RCA = combination of all OMI classes with RCA as the culprit vessel
- MI = combination of all MI classes
- NSTEMI = combination of all NSTEMI classes
- STEMI = combination of all STEMI classes

The ability to predict each individual OMI case – along with their location – was paramount in order to guide PCI operators to the culprit vessel in high urgency patients. Compared to the previous STEMI/NSTEMI study, predicting all MI cases not only has clinical applications but also helps evaluating model performance in test sets where only MI (yes/no) labels are available.

### Data splitting

Data were split into 70% training, 10% validation, and 20% test sets, where a patient could only be included in one of these sets. The test set was further divided equally into a random test set and a temporal test set, with the latter including patients whose first visit occurred in the final year (>=2016-01-01) of the study period. To augment the training data, multiple ECGs recorded at the same ED visit of a patient were used. In contrast, only one ECG per ED visit of a patient was used for validation and testing, and if multiple ECGs had been recorded at the ED visit for patients in the validation or test set, only one of these ECGs were retained at random.

### Model architecture and training

The model was based on a convolutional neural network (CNN) using a residual network (ResNet) architecture with skip connections, adapted for one-dimensional ECG signals. The model architecture has been described previously^16^, and the present study extends that previous model architecture. In addition to the ECGs, age and sex were used as predictors. Age and sex were first mapped to a higher-dimensional space (n=64), through a fully connected linear layer using ReLU activation to capture non-linear relationships. Age, sex, and ECGs were then concatenated before the final fully connected prediction layer. As in the STEMI/NSTEMI study, an ensemble of five independently trained model members was used, with their logits averaged to generate final predictions. The same hyperparameters as in the STEMI/NSTEMI study were used. Besides the weighted loss described below, the model architecture of the present study is identical to that of the STEMI/NSTEMI study.^12^

The model was trained using cross-entropy loss for the MI subclasses and binary cross-entropy loss was used for LBBB. These losses were combined as: weight_CE_*loss_CE_ + weight_BCE_*loss_BCE,_ where weight_CE_ +weight_BCE_ = 1. A hyperparameter search was conducted for determining the optimal BCE:CE weight ratio while all other hyperparameters were held fixed. The BCE:CE weight ratio of 0.3 yielded the best performance trade-off between MI subclass and LBBB classification (Supplementary Figure 12). The model loss was minimized over up to 150 epochs (Supplementary Figure 13).

### Model validation sets

The model was primarily evaluated in the SwED random and temporal test set described above, drawn from the same database used to train the model, including all outcomes which the model was trained to predict. Further, two external test sets (PTB-XL and CODE-II) including labels only for STEMI vs not STEMI were used to evaluate the model’s discriminatory performance for STEMIs.

The PTB-XL is a publicly available European database of 21,837 10-second 12-lead ECGs annotated with 71 different ECG statements, including cases of myocardial infarction.^17,18^ From PTB-XL, 63 acute myocardial infarction cases with a confirmed ST-elevation and 197 randomly selected controls without ST-elevation were manually curated from the PTB-XL database. The inclusion criteria of the PTB-XL test set used in this study has been described previously.^12^

The CODE-II dataset is an updated version of the first CODE dataset^16^, with exams collected and annotated using significant improvements in the internal operational system of the Telehealth Network of Minas Gerais (TNMG), Brazil. This system ensures standardized, high-quality ECG diagnostic classes. A complete description of the CODE-II dataset is under review and will be available soon at https://code.telessaude.hc.ufmg.br/. The dataset comprises 2,742,497 exams from 2,097,369 unique patients, recorded between January 2019 and December 2022. For this study, only the first exam from each patient was included, resulting in a total of 2,097,369 exams. The dataset primarily includes patients from primary care centers, along with some from hospitals, emergency departments, and ambulances. The median age is 54.2 years (IQR: 40.9, 66.3), with 40.9% of the patients being male. Among the 66 ECG CODE diagnostic classes, there is one comparable to STEMI (2,747 exams, 0.13%) and LBBB (40,490 exams, 1.93%) diagnostic classes, with the remaining classes aggregated as not STEMI (2,084,678 exams, 99.39%) for this study. Overlap is observed between exams labeled as LBBB with STEMI (56 exams) and not STEMI (30,485 exams).

To better understand the model’s classification errors in the CODE II dataset, we manually reviewed 20 ECGs that were misclassified by the model. This set included 10 false positives, defined as cases not labeled as STEMI where the model predicted a STEMI probability greater than 0.9, and 10 false negatives, where STEMI cases received a predicted probability less than 0.1. The ECGs were reviewed by a consultant cardiologist (GMMP), who was aware of the predicted probability.

### Metrics of discrimination and calibration

The C-statistic and average precision were calculated to evaluate the model’s discriminatory performance. These metrics were primarily assessed using one vs all other (OvA) comparisons, where each class was evaluated against all other classes combined, resulting in the ‘rest’ group being dominated by controls without myocarditis or pericarditis. Additionally, C-statistics were also computed using one vs one (OvO) comparisons, where each class was directly compared to another specific class.

As metrics of model calibration, the Brier score and the Expected Calibration Error (ECE) were calculated. The Brier score was calculated as the mean squared error on the probability scale, both in binary OvA calculation but also as multiclass calculation, averaging the OvA Brier scores across all classes. The Expected Calibration Error (ECE) was computed as the weighted average of the absolute differences between the accuracy and the predicted confidence within 10 probability bins.

Confidence for each prediction was defined as the maximum predicted probability across classes, reflecting the model’s certainty in its top choice.

We visualized the calibration of our model using binned calibration plots of the observed case frequency vs the average predicted probability. Given the low number of cases for some of the outcomes, which can result in very few cases in a bin, we used the combined random and temporal test set for these plots to increase the total number of cases. Up to 20 bins were used, defined based on quantile cutoffs of the predicted probabilities of the evaluated outcome class ensuring that at least 10 cases of the given outcome were present in a given bin.

### Model performance in subgroups

We evaluated whether the model made incorrect predictions (with a high probability) for any of the recorded comorbidities of the patients. For each outcome class, we selected all records with a predicted probability of the class >0.5. Among these, we compared correctly versus incorrectly classified patients based on the true class labels. A non-parametric conditional independence test based on the maximum test statistic was then used to assess whether specific 3-character ICD-10 diagnosis codes were over-or underrepresented among the misclassified patients vs correctly classified patients. All available 3-character ICD-10 codes were tested, and those with a false discovery rate (FDR) < 0.01 were highlighted.

In a manually selected list of subgroups, where the model performance might differ, we calculated the C-statistic in OvA comparisons. Given that some subgroups are small, we used the combined random and temporal test set in SwED for the groups control, nOMI, and OMI.

