## Supplementary material for "A deep learning ECG model for localization of occlusion myocardial infarction": All supplementary materials

### Supplement

**Supplementary Table 1.** Number of ECGs per outcome class and set in the SwED study sample. The percent out of all ECGs in the given set/column is provided within parentheses. OMI, occlusion MI; STEMI, ST-elevation myocardial infarction; NSTEMI, non-ST-elevation myocardial infarction; LM/LAD, left main coronary artery+left anterior descending artery; LCX, left circumflex artery; RCA, right coronary artery; Test (rand), random test set; Test (temp), temporal test set.

| | **Training** | **Validation** | **Test (rand)** | **Test (temp)** |
|---|---|---|---|---|
| **Control** | 410,434 (98.88) | 48,699 (99.16) | 48,680 (99.10) | 26,697 (98.68) |
| Without perimyocarditis | 409,303 (98.61) | 48,590 (98.94) | 48,566 (98.87) | 26,606 (98.35) |
| With perimyocarditis | 1,131 (0.27) | 109 (0.22) | 114 (0.23) | 91 (0.34) |
| **nOMI** | 3,416 (0.82) | 296 (0.60) | 339 (0.69) | 228 (0.84) |
| NSTEMI | 2,812 (0.68) | 248 (0.50) | 278 (0.57) | 187 (0.69) |
| STEMI | 604 (0.15) | 48 (0.10) | 61 (0.12) | 41 (0.15) |
| **OMI** | 1,236 (0.30) | 115 (0.23) | 104 (0.21) | 128 (0.47) |
| NSTEMI, LM/LAD | 93 (0.02) | 20 (0.04) | 7 (0.01) | 18 (0.07) |
| NSTEMI, LCX | 198 (0.05) | 11 (0.02) | 10 (0.02) | 10 (0.04) |
| NSTEMI, RCA | 157 (0.04) | 10 (0.02) | 11 (0.02) | 7 (0.03) |
| STEMI, LM/LAD | 346 (0.08) | 37 (0.08) | 34 (0.07) | 40 (0.15) |
| STEMI, LCX | 99 (0.02) | 10 (0.02) | 7 (0.01) | 14 (0.05) |
| STEMI, RCA | 343 (0.08) | 27 (0.05) | 35 (0.07) | 39 (0.14) |

**Supplementary Table 2**. Performance metrics of the model in the two test sets in SwED. Sample size is given by each testset and outcome label together with percent out of the given testset in Supplementary Table 1. Performance metrics are given as median (minimum-maximum) over ten trained models initiated with different seeds; each of the ten models is an ensemble consisting of five model members. Arrows indicate direction of better performance. We compute the metrics as class *vs.* all other (OvA). For the Brier score and ECE a multi-class calculation is provided, calculated for the ten MI subclasses. MI, myocardial infarction; OMI, occlusion MI; STEMI, ST-elevation myocardial infarction; NSTEMI, non-ST-elevation myocardial infarction; LM/LAD, left main coronary artery+left anterior descending artery; LCX, left circumflex artery; RCA, right coronary artery; AP, Average Precision or equivalently Area Under the Precision-Recall curve; ECE, Expected Calibration Error; Test (rand), random test set; Test (temp), temporal test set.

| **Metric** | **Outcome** | **Test (rand)** | **Test (temp)** |
|---|---|---|---|
| C-statistic (↑) | Control, without perimyocarditis | 0.8762 (0.8724,0.8784) | 0.8906 (0.8869,0.8936) |
| | Control, with perimyocarditis | 0.9078 (0.8980,0.9078) | 0.8696 (0.8650,0.8745) |
| | nOMI, NSTEMI | 0.8542 (0.8531,0.8584) | 0.8696 (0.8629,0.8717) |
| | OMI, NSTEMI, LM/LAD | 0.9488 (0.9474,0.9653) | 0.8940 (0.8869,0.9068) |
| | OMI, NSTEMI, LCX | 0.8320 (0.8130,0.8438) | 0.9782 (0.9630,0.9782) |
| | OMI, NSTEMI, RCA | 0.9122 (0.8872,0.9202) | 0.9945 (0.9935,0.9963) |
| | nOMI, STEMI | 0.9467 (0.9463,0.9541) | 0.9593 (0.9534,0.9593) |
| | OMI, STEMI, LM/LAD | 0.9768 (0.9768,0.9858) | 0.9926 (0.9908,0.9931) |
| | OMI, STEMI, LCX | 0.9648 (0.9591,0.9697) | 0.9949 (0.9940,0.9957) |
| | OMI, STEMI, RCA | 0.9792 (0.9775,0.9824) | 0.9866 (0.9866,0.9918) |
| | LBBB | 0.9855 (0.9841,0.9855) | 0.9903 (0.9896,0.9904) |
| | MI | 0.8907 (0.8895,0.8943) | 0.9206 (0.9161,0.9220) |
| | OMI | 0.9534 (0.9514,0.9571) | 0.9764 (0.9756,0.9796) |
| | OMI LM/LAD | 0.9712 (0.9712,0.9794) | 0.9617 (0.9585,0.9661) |
| | OMI LCX | 0.8851 (0.8728,0.8902) | 0.9856 (0.9786,0.9858) |
| | OMI RCA | 0.9636 (0.9571,0.9671) | 0.9877 (0.9877,0.9922) |
| | nOMI | 0.8699 (0.8689,0.8737) | 0.8846 (0.8780,0.8858) |
| | STEMI | 0.9638 (0.9638,0.9704) | 0.9819 (0.9812,0.9834) |
| | NSTEMI | 0.8580 (0.8563,0.8621) | 0.8827 (0.8762,0.8844) |
| AP (↑) | Control, without perimyocarditis | 0.9978 (0.9977,0.9978) | 0.9970 (0.9969,0.9971) |
| | Control, with perimyocarditis | 0.2252 (0.2174,0.2309) | 0.2329 (0.2173,0.2348) |
| | nOMI, NSTEMI | 0.1068 (0.1024,0.1119) | 0.1661 (0.1515,0.1667) |
| | OMI, NSTEMI, LM/LAD | 0.0094 (0.0071,0.0237) | 0.0411 (0.0334,0.0937) |
| | OMI, NSTEMI, LCX | 0.0019 (0.0014,0.0021) | 0.1668 (0.0635,0.2742) |
| | OMI, NSTEMI, RCA | 0.0963 (0.0963,0.2283) | 0.0323 (0.0317,0.0455) |
| | nOMI, STEMI | 0.1703 (0.1560,0.1944) | 0.1304 (0.1261,0.1385) |
| | OMI, STEMI, LM/LAD | 0.5391 (0.5313,0.5812) | 0.5650 (0.5355,0.5945) |
| | OMI, STEMI, LCX | 0.1995 (0.1995,0.2787) | 0.2756 (0.2634,0.3071) |
| | OMI, STEMI, RCA | 0.5437 (0.5285,0.5601) | 0.5919 (0.5886,0.6212) |
| | LBBB | 0.8755 (0.8692,0.8755) | 0.9229 (0.9183,0.9237) |
| | MI | 0.3681 (0.3593,0.3732) | 0.5383 (0.5247,0.5390) |
| | OMI | 0.4975 (0.4802,0.4975) | 0.5892 (0.5820,0.5950) |
| | OMI LM/LAD | 0.5245 (0.5110,0.5460) | 0.4827 (0.4581,0.5041) |
| | OMI LCX | 0.0890 (0.0745,0.1499) | 0.2757 (0.2426,0.3354) |

| | | | |
|---|---|---|---|
| | OMI RCA | 0.5319 (0.5061,0.5411) | 0.6012 (0.5919,0.6336) |
| | nOMI | 0.1695 (0.1611,0.1760) | 0.2057 (0.1942,0.2082) |
| | STEMI | 0.5795 (0.5699,0.5848) | 0.6861 (0.6662,0.6861) |
| | NSTEMI | 0.1252 (0.1205,0.1287) | 0.2206 (0.1957,0.2206) |
| Brier **(↓)** | Control, without perimyocarditis | 0.0088 (0.0087,0.0090) | 0.0109 (0.0109,0.0112) |
| | Control, with perimyocarditis | 0.0021 (0.0020,0.0021) | 0.0029 (0.0029,0.0029) |
| | nOMI, NSTEMI | 0.0054 (0.0054,0.0054) | 0.0063 (0.0063,0.0063) |
| | OMI, NSTEMI, LM/LAD | 0.0001 (0.0001,0.0001) | 0.0007 (0.0007,0.0007) |
| | OMI, NSTEMI, LCX | 0.0002 (0.0002,0.0002) | 0.0003 (0.0003,0.0004) |
| | OMI, NSTEMI, RCA | 0.0002 (0.0002,0.0002) | 0.0003 (0.0003,0.0003) |
| | nOMI, STEMI | 0.0011 (0.0011,0.0011) | 0.0014 (0.0014,0.0014) |
| | OMI, STEMI, LM/LAD | 0.0004 (0.0004,0.0004) | 0.0008 (0.0008,0.0009) |
| | OMI, STEMI, LCX | 0.0001 (0.0001,0.0001) | 0.0005 (0.0005,0.0005) |
| | OMI, STEMI, RCA | 0.0004 (0.0004,0.0005) | 0.0008 (0.0008,0.0009) |
| | LBBB | 0.0090 (0.0090,0.0092) | 0.0029 (0.0029,0.0030) |
| | MI | 0.0069 (0.0068,0.0071) | 0.0083 (0.0083,0.0085) |
| | OMI | 0.0014 (0.0014,0.0014) | 0.0028 (0.0028,0.0028) |
| | OMI LM/LAD | 0.0005 (0.0005,0.0005) | 0.0014 (0.0014,0.0014) |
| | OMI LCX | 0.0003 (0.0003,0.0003) | 0.0008 (0.0008,0.0008) |
| | OMI RCA | 0.0006 (0.0006,0.0006) | 0.0010 (0.0009,0.0010) |
| | nOMI | 0.0063 (0.0062,0.0064) | 0.0074 (0.0074,0.0075) |
| | STEMI | 0.0016 (0.0016,0.0017) | 0.0025 (0.0025,0.0026) |
| | NSTEMI | 0.0059 (0.0058,0.0059) | 0.0072 (0.0072,0.0073) |
| | Multi | 0.0189 (0.0187,0.0191) | 0.0249 (0.0249,0.0253) |
| ECE **(↓)** | Multi | 0.0106 (0.0105,0.0108) | 0.0143 (0.0143,0.0147) |

**Supplementary Table 3.** Metrics of discriminatory performance in the external validation tests CODE-II (Brazilian) and PTB-XL (European). Only labels for the presence of STEMI (yes/no) are available in these test sets and can be evaluated. Arrows indicate the direction of better performance. The C-statistic and average precision (AP) are provided.

| **Metric** | **Outcome** | **CODE-II** | **PTB-XL** |
|---|---|---|---|
| C-statistic **(↑)** | STEMI | 0.9871 | 0.9980 |
| | Not STEMI | 0.8798 | 0.9980 |
| | LBBB | 0.9870 | NA |
| AP **(↑)** | STEMI | 0.5396 | 0.9936 |
| | Not STEMI | 0.9991 | 0.9993 |
| | LBBB | 0.8732 | NA |

**Supplementary Table 4.** Over-/underrepresented diagnoses when comparing correct classifications with misclassifications for a given observed class, restricted to ECGs with a high predicted probability (Pr>0.5). Tested in an asymptotic general independence test with a two-sided alternative hypothesis. All diagnoses at time of the visit are included (all diagnosis positions). Three-character ICD10 codes were tested unless only the ICD10 block was available in the data. All results with a false discovery rate (Benjamini-Hochberg) < 0.01 are reported.

| Truth | Misclassification | Over/under-represented diagnosis | OR |
|---|---|---|---|
| Control | nOMI | X70 | 335.0 |
| Control | nOMI | T71 | 297.2 |
| Control | nOMI | N99 | 245.0 |
| Control | nOMI | K04 | 191.4 |
| Control | nOMI | K27 | 178.4 |
| Control | OMI | I51 | 121.2 |
| Control | nOMI | J95 | 111.9 |
| Control | OMI | I46 | 81.6 |
| Control | OMI | I40 | 79.9 |
| Control | nOMI | I33 | 74.6 |
| Control | nOMI | U99 | 61.0 |
| Control | nOMI | K55 | 51.7 |
| Control | nOMI | M13 | 48.9 |
| Control | nOMI | Z33 | 48.9 |
| Control | nOMI | I46 | 42.5 |
| Control | OMI | F31 | 34.3 |
| Control | nOMI | A40 | 32.3 |
| Control | nOMI | I60 | 32.3 |
| Control | nOMI | I40 | 28.6 |
| Control | nOMI | I67 | 28.0 |
| Control | nOMI | Y88 | 26.9 |
| Control | OMI | I42 | 26.2 |
| Control | nOMI | D86 | 25.3 |
| Control | OMI | K21 | 24.1 |
| Control | OMI | R47-R49 | 24.1 |
| Control | nOMI | F60 | 21.6 |
| Control | OMI | I47 | 19.0 |
| Control | nOMI | N17 | 17.8 |
| Control | nOMI | A41 | 13.3 |
| Control | nOMI | I25 | 10.6 |
| Control | nOMI | J15 | 8.3 |
| Control | nOMI | N18 | 7.7 |
| Control | nOMI | I20 | 6.2 |
| Control | nOMI | I50 | 5.3 |
| Control | nOMI | E11 | 5.0 |

**Supplementary Table 5**. Manual review of 20 misclassified ECGs. The upper section presents 10 false positives (Scenario 1), and the lower section shows 10 false negatives (Scenario 2). Diagnoses were reassigned by a consultant cardiologist.

| **Scenario 1 - False Positives: Model predicted STEMI (probability > 0.9), originally labeled as Not STEMI** | | | | | |
|---|---|---|---|---|---|
| Predicted probability | | | | | Consultant Cardiologist Review |
| nOMI, STEMI | OMI, STEMI, LM/LAD | OMI, STEMI, LCX | OMI, STEMI, RCA | STEMI | |
| 0.24 | 0.01 | 0.02 | 0.70 | 0.97 | Left atrial enlargement, STEMI |
| 0.42 | 0.54 | 0.00 | 0.01 | 0.97 | STEMI |
| 0.07 | 0.00 | 0.02 | 0.90 | 0.99 | STEMI |
| 0.34 | 0.54 | 0.01 | 0.01 | 0.90 | ST elevation: pericarditis pattern |
| 0.20 | 0.80 | 0.00 | 0.00 | 1.00 | STEMI |
| 0.20 | 0.78 | 0.00 | 0.00 | 0.98 | Q wave pathological, STEMI |
| 0.20 | 0.00 | 0.07 | 0.66 | 0.93 | STEMI |
| 0.28 | 0.63 | 0.00 | 0.01 | 0.92 | STEMI |
| 0.25 | 0.66 | 0.00 | 0.01 | 0.92 | First degree AVB, left atrial enlargement, pathological Q wave, ST elevation (dyskinetic area), nonspecific intraventricular conduction delay |
| 0.16 | 0.01 | 0.04 | 0.75 | 0.96 | RBBB, STEMI |
| **Scenario 2 - False Negatives: Model predicted Not STEMI (probability < 0.1), originally labeled as STEMI** | | | | | |
| Predicted probability | | | | | Consultant Cardiologist Review |
| nOMI, STEMI | OMI, STEMI, LM/LAD | OMI, STEMI, LCX | OMI, STEMI, RCA | STEMI | |
| 0.01 | 0.00 | 0.00 | 0.00 | 0.01 | Left atrial enlargement, left anterior hemiblock, left ventricular hypertrophy,STEMI, PVC |
| 0.01 | 0.01 | 0.00 | 0.00 | 0.02 | Technical problem |
| 0.00 | 0.00 | 0.00 | 0.00 | 0.01 | ST elevation: pericarditis pattern |
| 0.00 | 0.00 | 0.00 | 0.00 | 0.00 | Sinus tachycardia, STEMI |
| 0.00 | 0.00 | 0.00 | 0.00 | 0.00 | Left anterior hemiblock, left atrial enlargement, left ventricular hypertrophy, technical problem |
| 0.02 | 0.01 | 0.00 | 0.01 | 0.05 | AF, STEMI |
| 0.00 | 0.00 | 0.00 | 0.00 | 0.00 | Left atrial enlargement, LVH, PVC |
| 0.02 | 0.02 | 0.00 | 0.00 | 0.05 | STEMI, Subendocardial ischemia |
| 0.00 | 0.00 | 0.00 | 0.00 | 0.00 | LBBB, STEMI |
| 0.01 | 0.01 | 0.00 | 0.00 | 0.01 | Technical problem |

**Supplementary Figure 1**

**Panel A)** Inclusion and exclusion criteria for the main study sample (the Swedish Emergency Department database [SwED]).

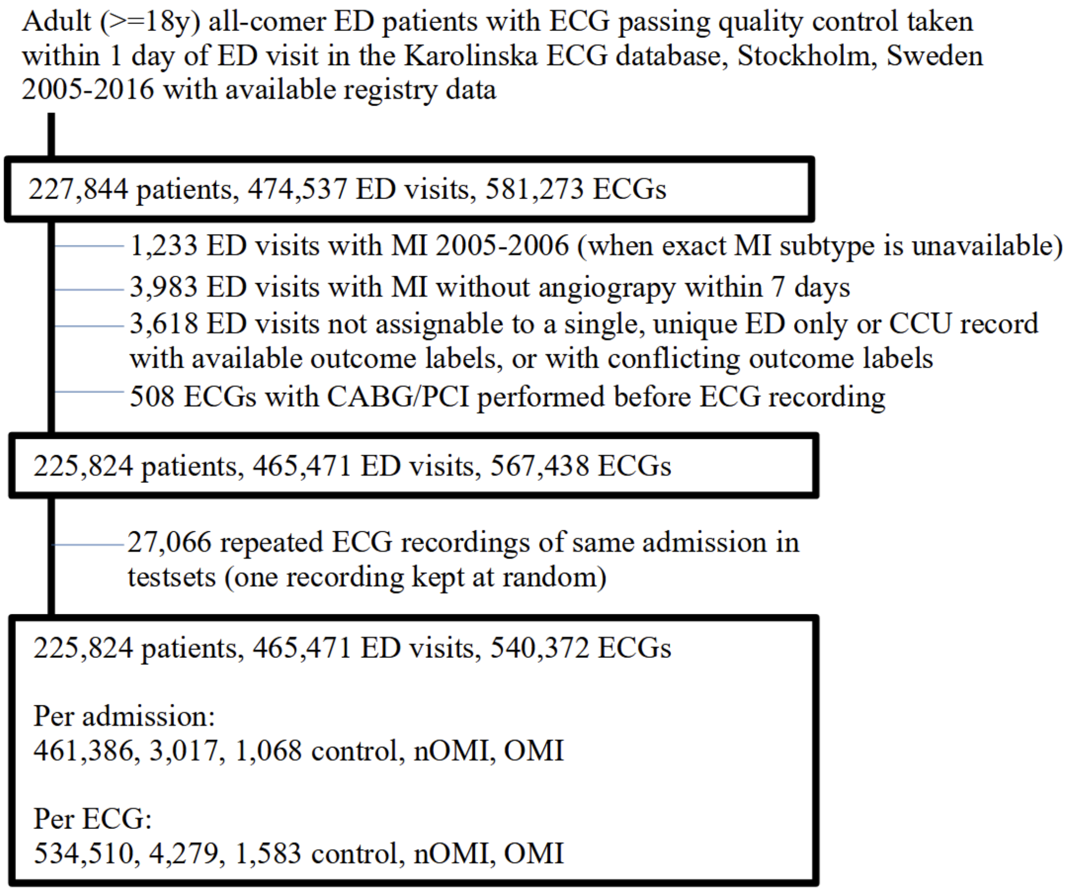


**Panel B)** Data splits of the derived main study sample.

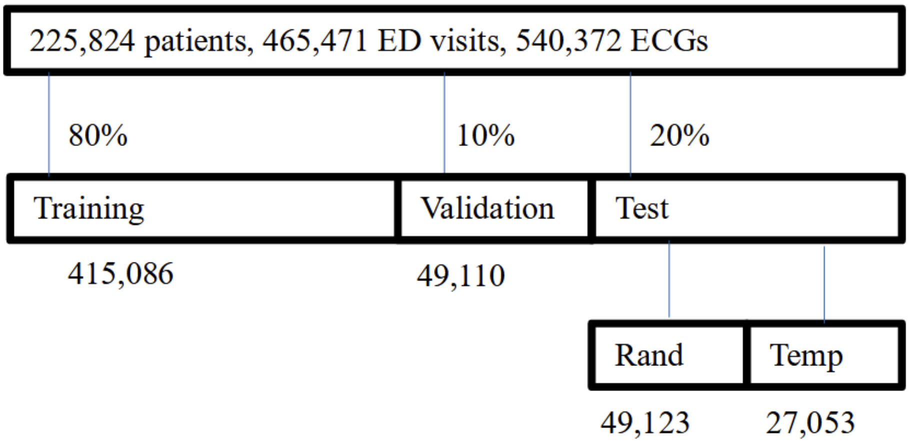

**Supplementary Figure 2**. Hierarchical organization of outcome classes used in this study. All MI and control subclasses are mutually exclusive. LBBB is a co-diagnostic class that may co-occur with any other subclass from the MI or control categories. Abbreviations: MI, myocardial infarction; OMI, occlusion MI; nOMI, non-occlusion MI; STEMI, ST-elevation MI; NSTEMI, non-ST-elevation MI; LM/LAD, left main coronary artery and left anterior descending artery; LCX, left circumflex artery; RCA, right coronary artery; LBBB, left bundle-branch block.

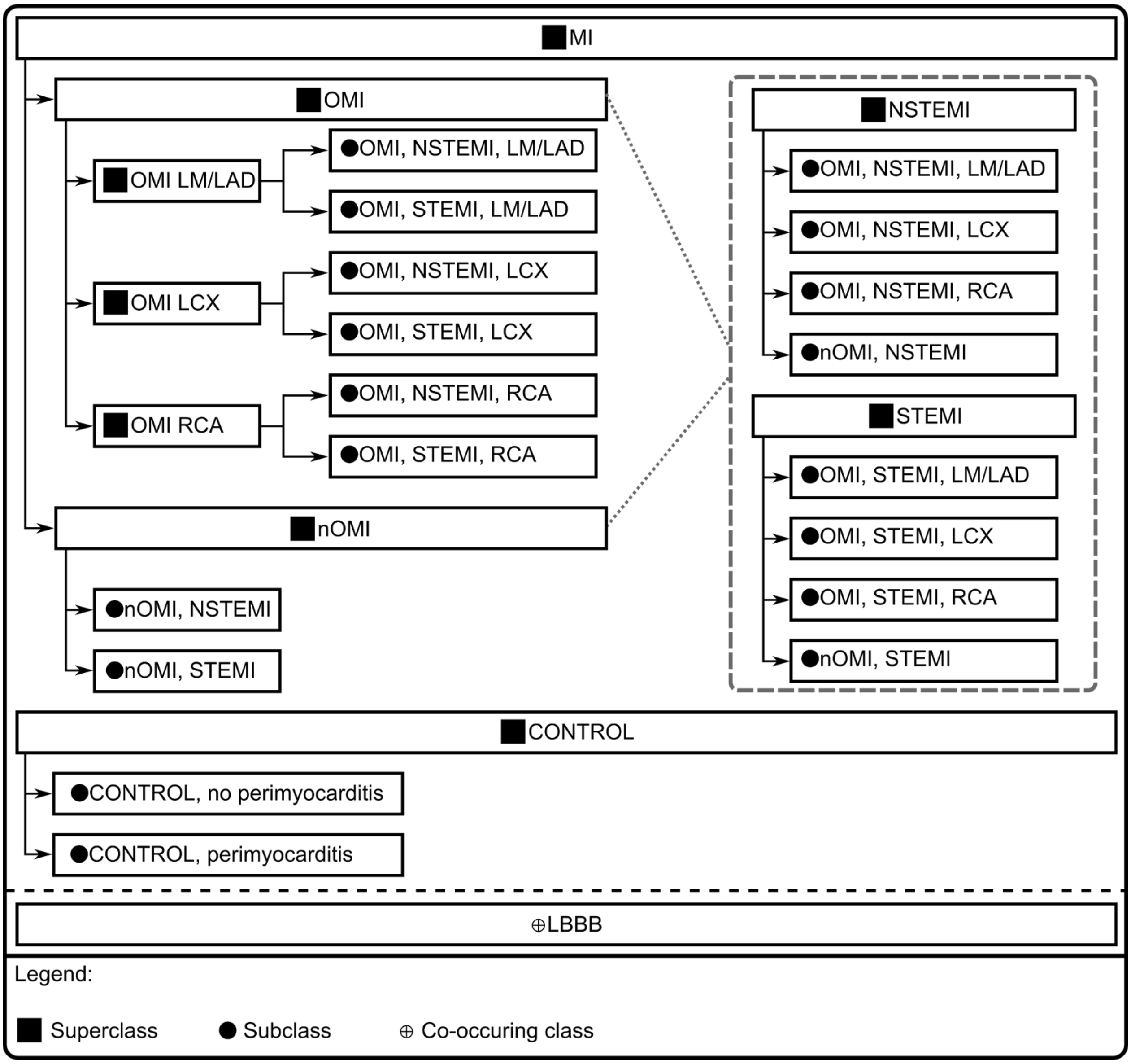

**Supplementary Figure 3.** Number of ECGs per year with separate panels for myocardial infarction (MI) with an urgent occlusion (OMI), MI without OMI (nOMI), and controls free from MI. MI cases from 2005-2006 were excluded due to missing database annotations needed for subcategorization as highlighted by the dotted vertical line. The dashed line represents the cutoff for the temporal test set.

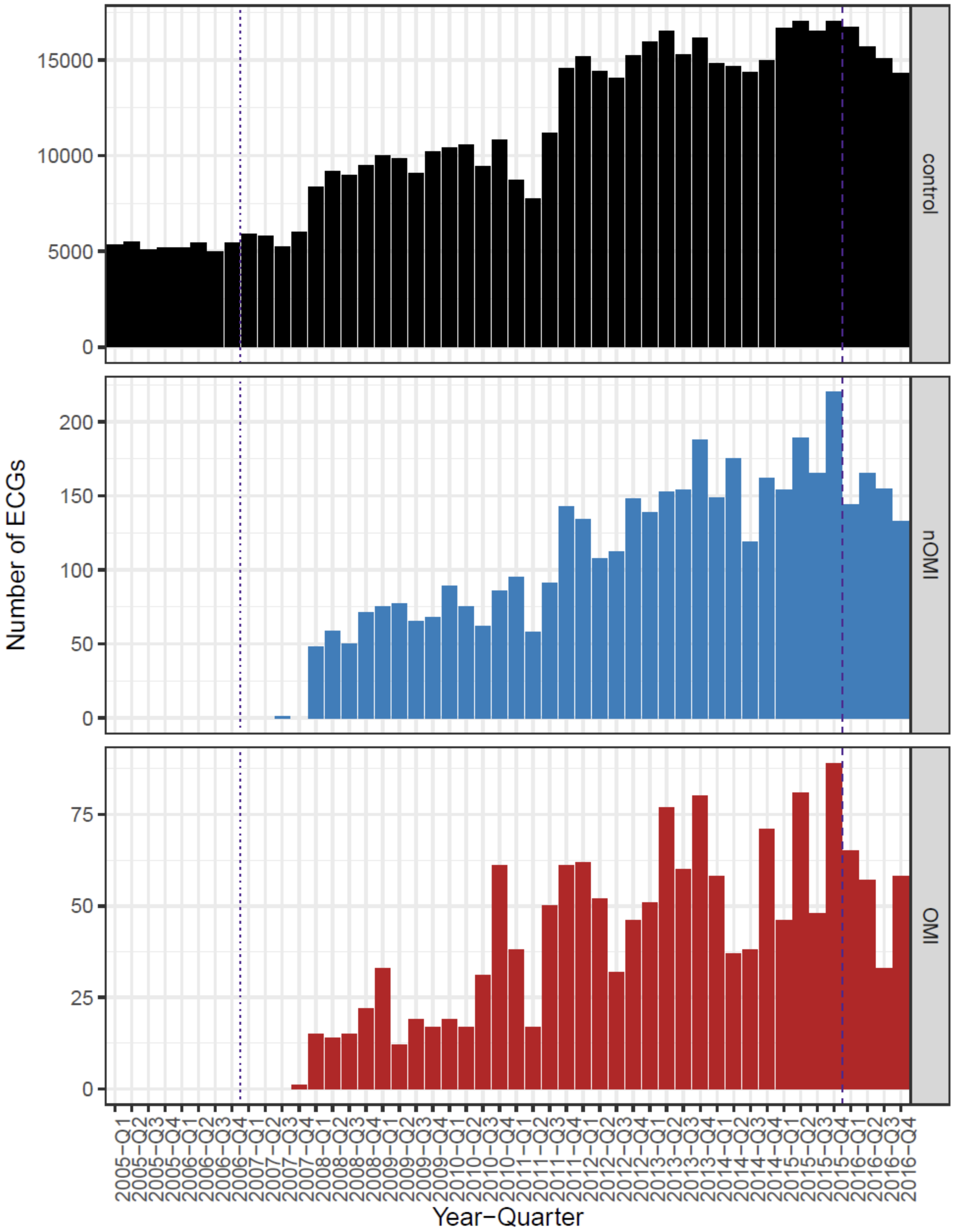

**Supplementary Figure 4.** Number of ECGs per year by outcome label, with separate panels for controls, MI, and LBBB. LBBB can co-occur with other classes. MI cases from 2005-2006 were excluded due to missing database annotations needed for subcategorization as highlighted by the dotted vertical line. The dashed line represents the cutoff for the temporal test set.

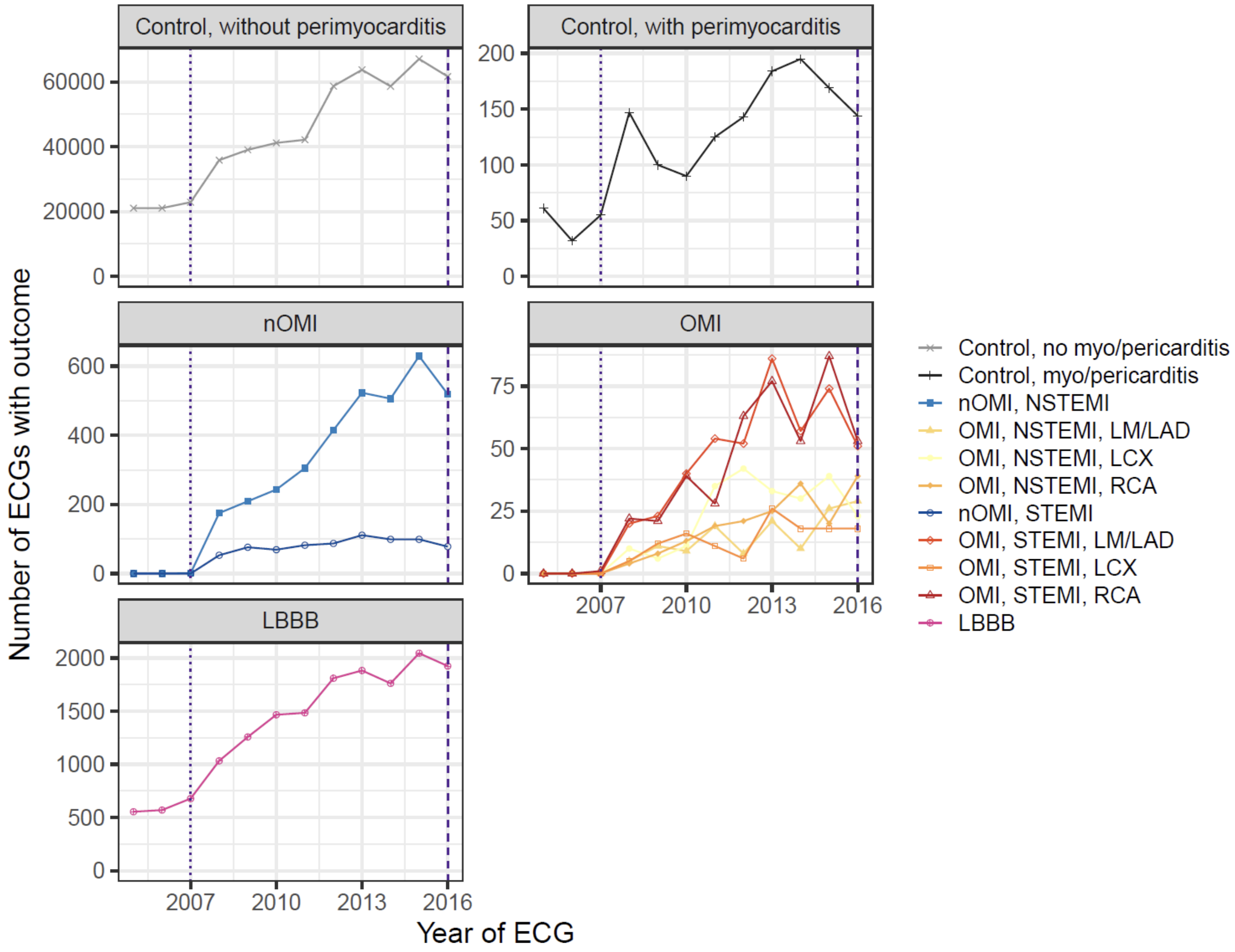

**Supplementary Figure 5.** Kaplan-Meier analysis of time to percutaneous coronary intervention (PCI) following the patients for up to 30 days after ED admission. Stratified by ED year tertiles and OMI.

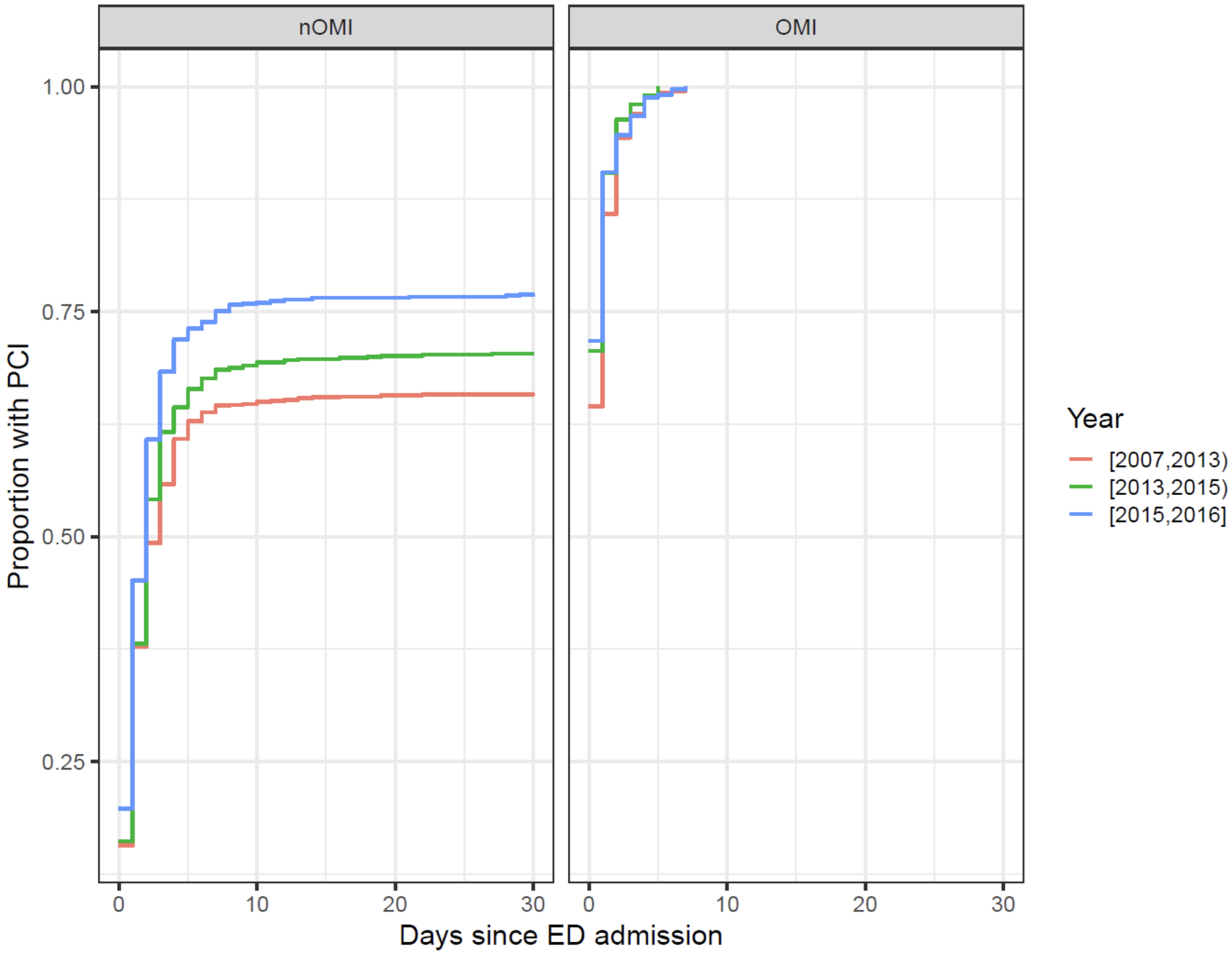

**Supplementary Figure 6.** Discriminative performance (C-statistic) when comparing a given class (x-axis) with all other classes in the random or temporal test set. All predicted sub classes of the model are included. Error bars represent min/max C-statistic from the model initiated with 10 different random seeds. The dashed horizontal line represents a random guess.

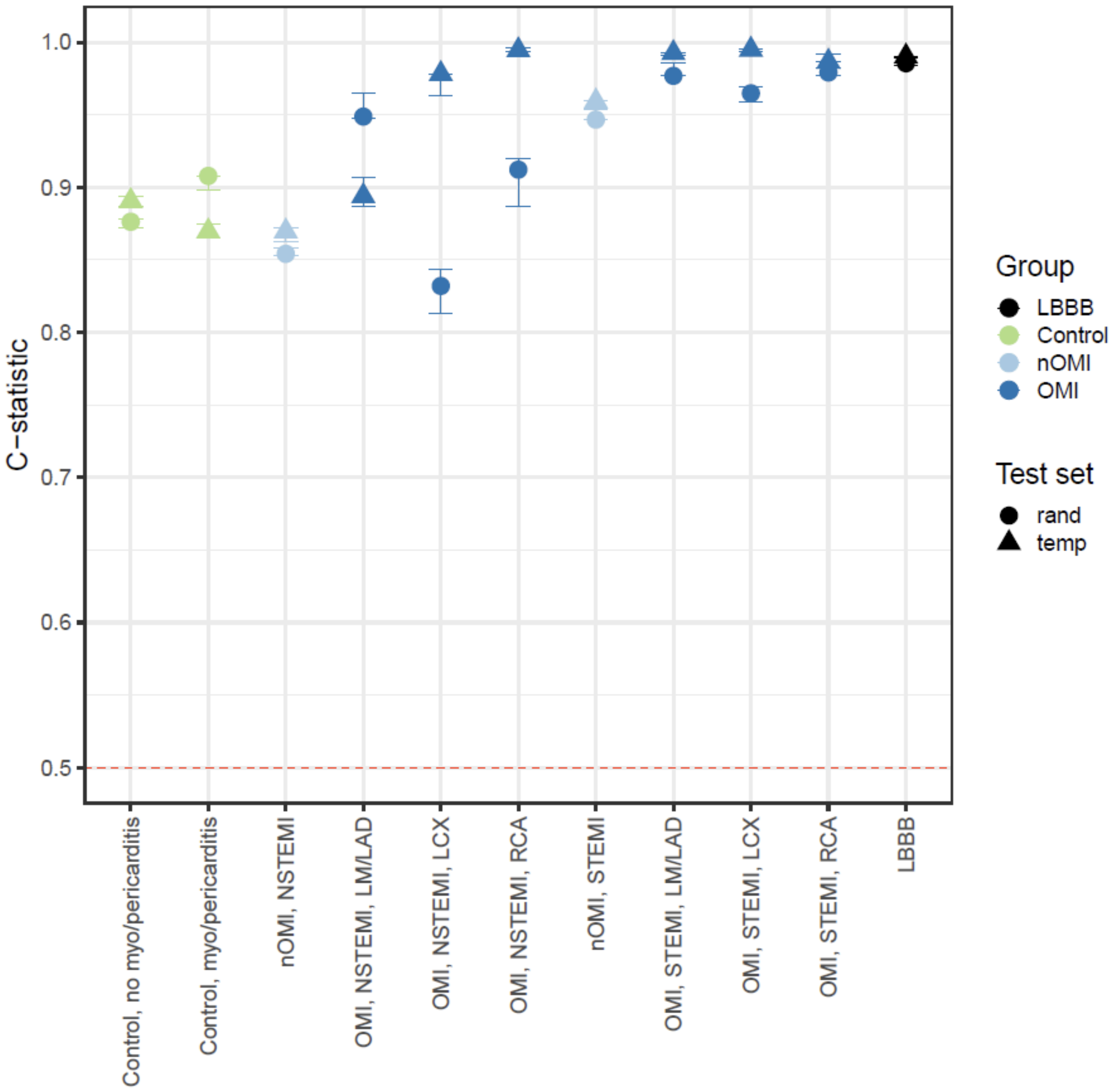

**Supplementary Figure 7.** Panel A showing receiver operating characteristics (ROC) curves, separately for each test set. Panel B showing precision-recall (PR) curves of each outcome (one versus rest), calculated from the combined test sets, using 50 fixed, evenly spaced bins, using linear interpolation between the points to avoid extremely jagged curves due to few cases in each outcome class.

**Panel A)**

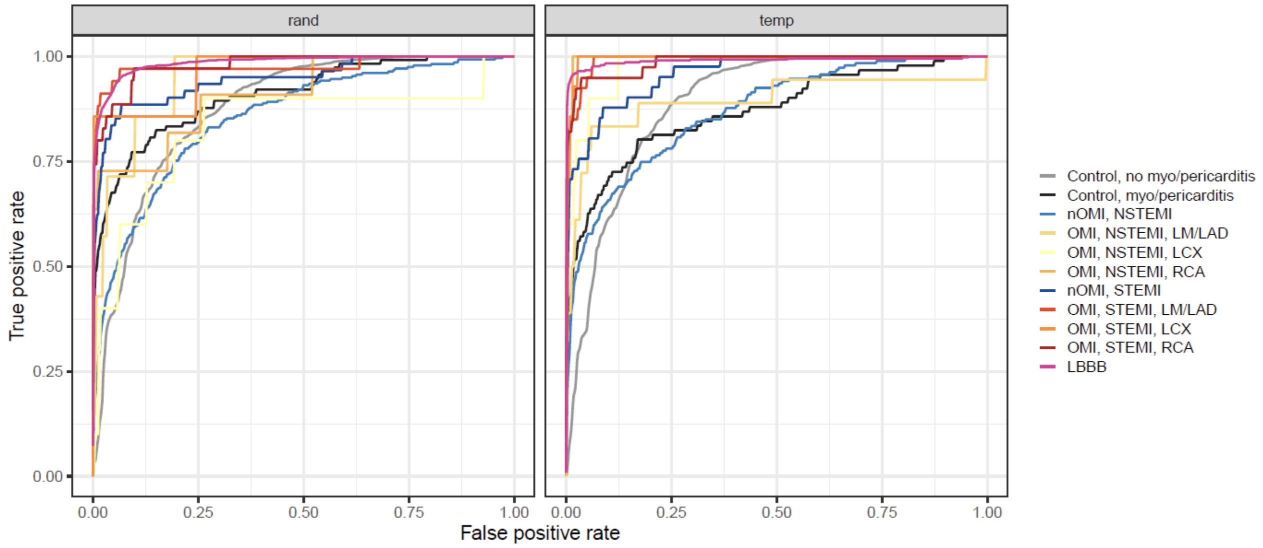


**Panel B)**

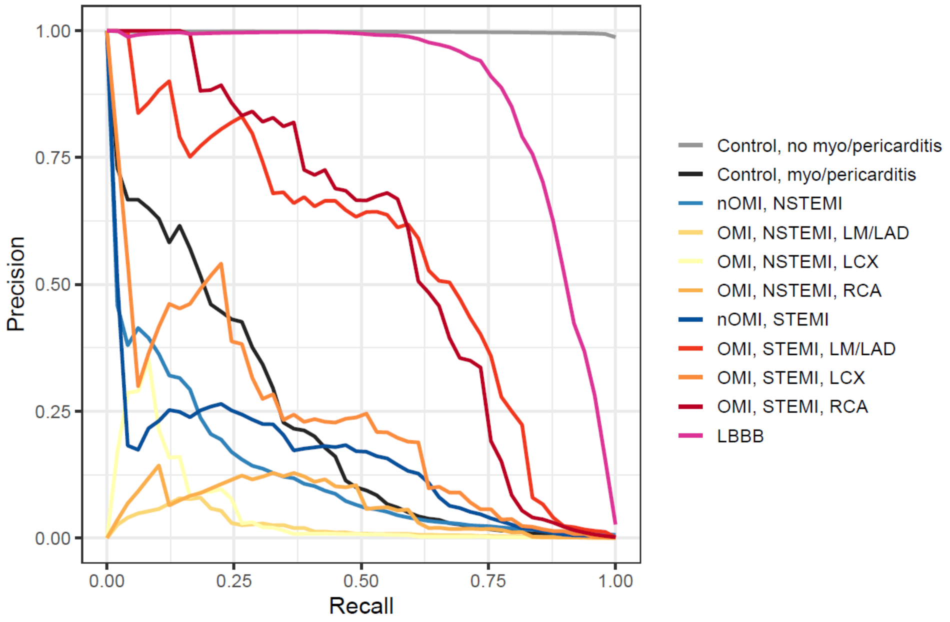

**Supplementary Figure 8.** Discriminative performance (C-statistic) in one versus one (OvO) comparisons. The predicted probability and truth label of outcome 2 (y-axis) is used in the calculation.

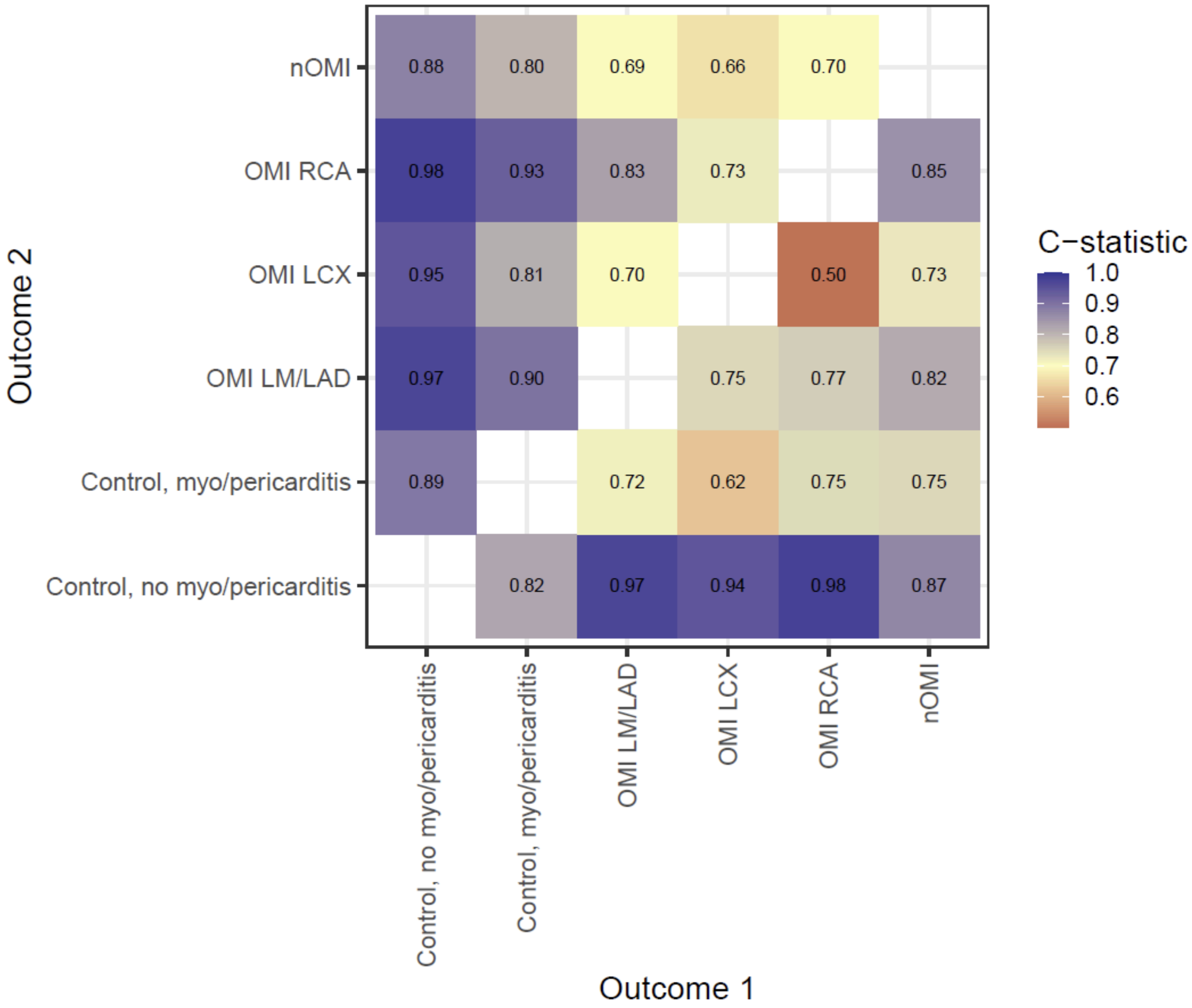

**Supplementary Figure 9.** Binned calibration plots in both SwED tests combined. Up to 20 quantile bins included with the requirement of at least 10 cases were present in each bin. The binning resolution is sometimes poor due to few cases with the given outcome class.

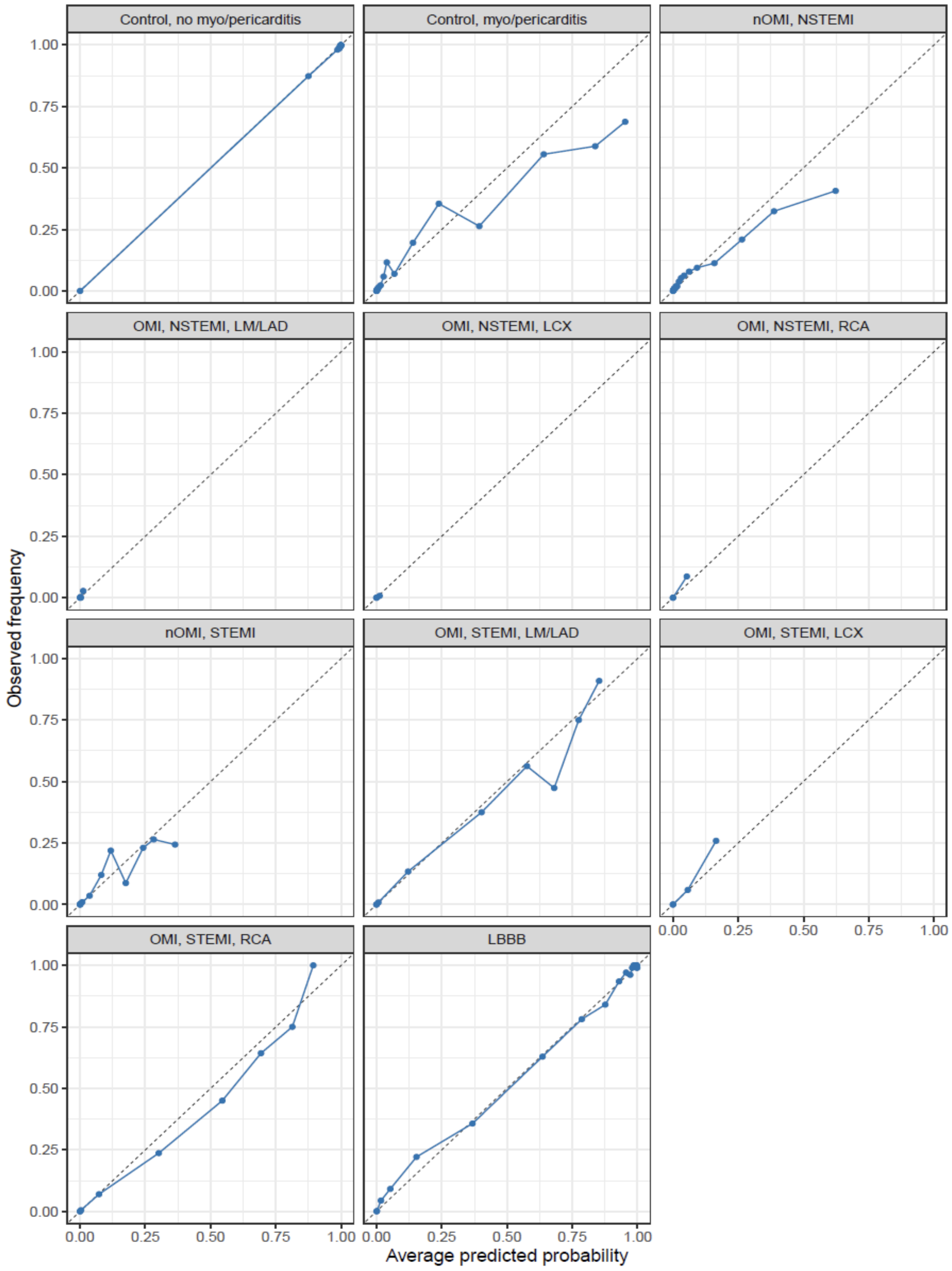

**Supplementary Figure 10.** Discriminative performance (C-statistic) when comparing a given class (color) with all other classes in the both SwED test sets combined, stratified by demographics, comorbidities, and technical factors, including age tertiles, sex, did the patient visit the emergency department at Karolinska Hospital (main source of data) or another emergency department in the Stockholm region, ECGs recorded using the most common machine type (MAC55) or not, ECGs recorded using the most common software (v237) or not, patient with a prevalent hospitalisation due to cardiovascular disease prior to the ED visit, patients with prevalent LBBB.

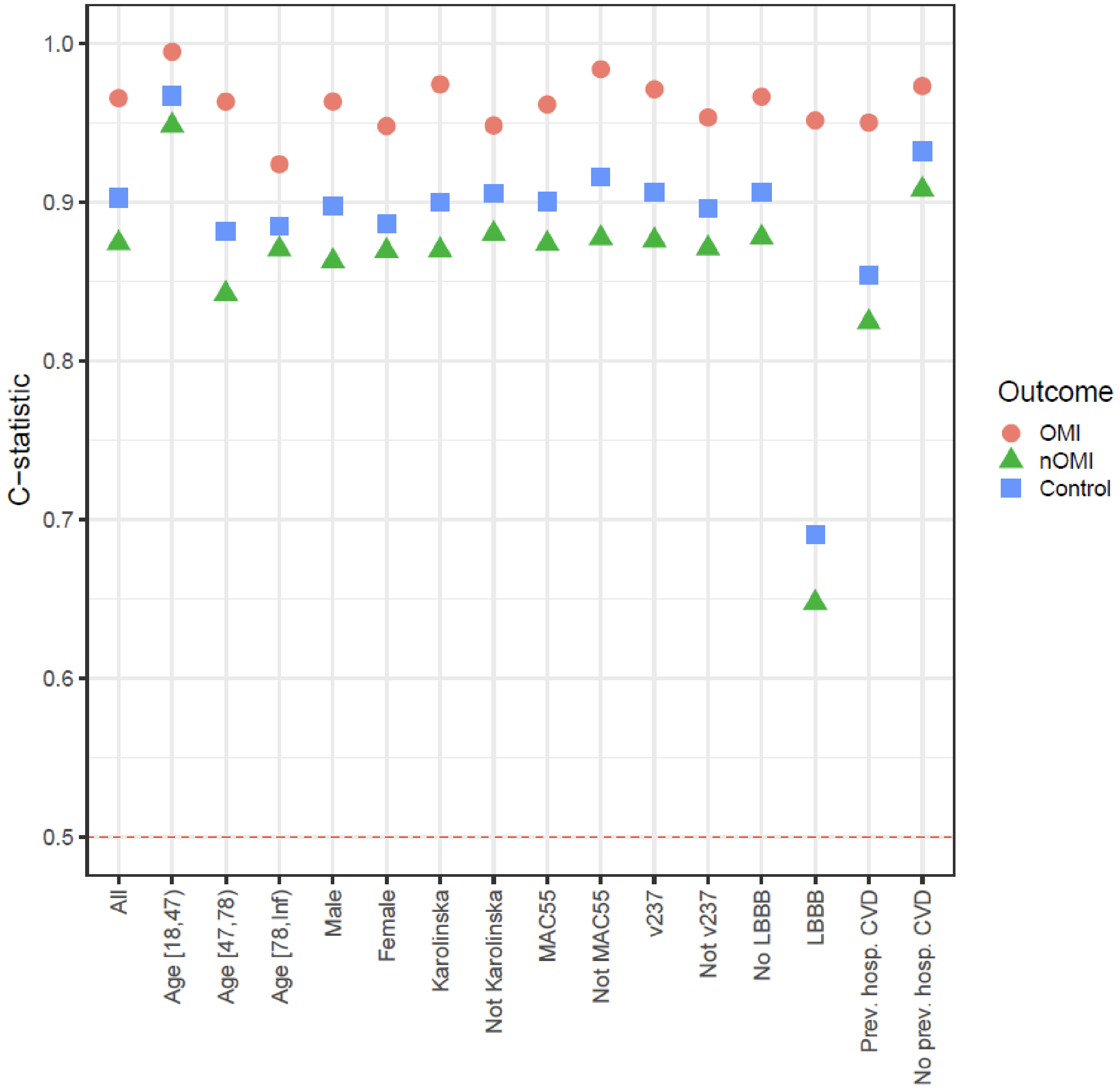

**Supplementary Figure 11.** Predicted probabilities of left bundle branch block (LBBB) from a machine learning model trained on Brazilian ECGs[15]. Separate panels for those with or without a prevalent diagnosis of LBBB in the SwED data. The dashed vertical line marks the lower cutoff (>=0.5) for classifying an ECG as LBBB. Note that all patients with prevalent LBBB will not have a diagnosis in the data (LBBB unknown/missed or diagnosis code not set). X-axis is on a log10 scale.

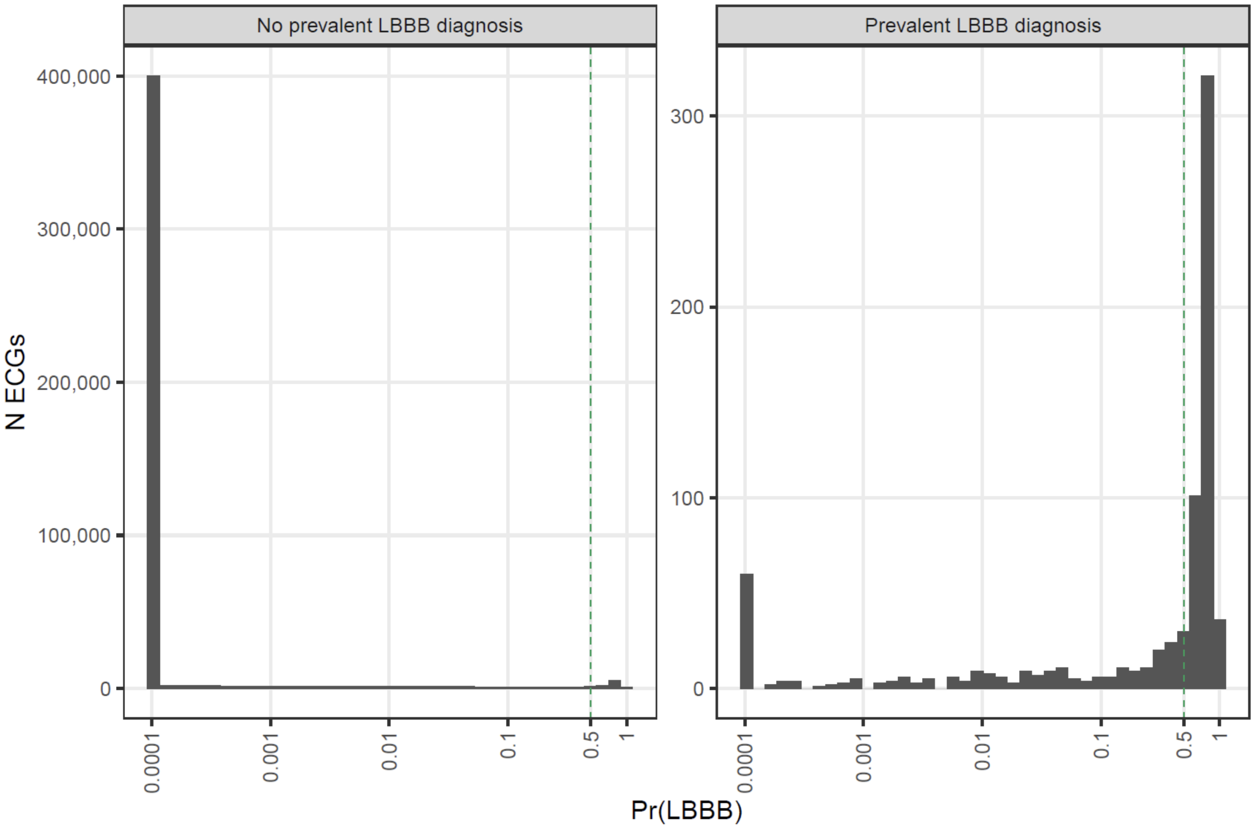

**Supplemental Figure 12.** Evaluation metrics were calculated in the validation set, for separate model fits with the BCE:CE weight ratio hyperparameter ranging from 0.05 to 20. Panel A: The C-statistic (reflecting model discrimination) was calculated in one-vs-rest for each of the 11 classes. Panel B: The Brier score (reflecting both the model discrimination and calibration) was calculated for LBBB (1 class) and the multi-class MI outcome (10 classes)

**Panel A**

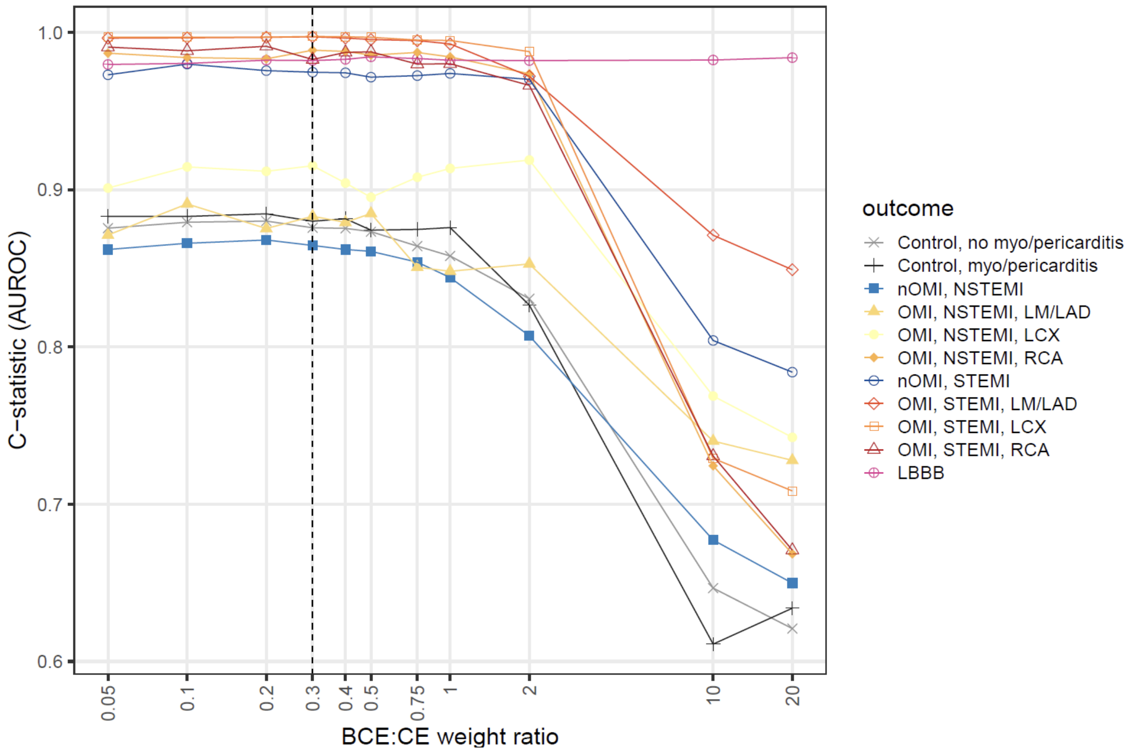


**Panel B**

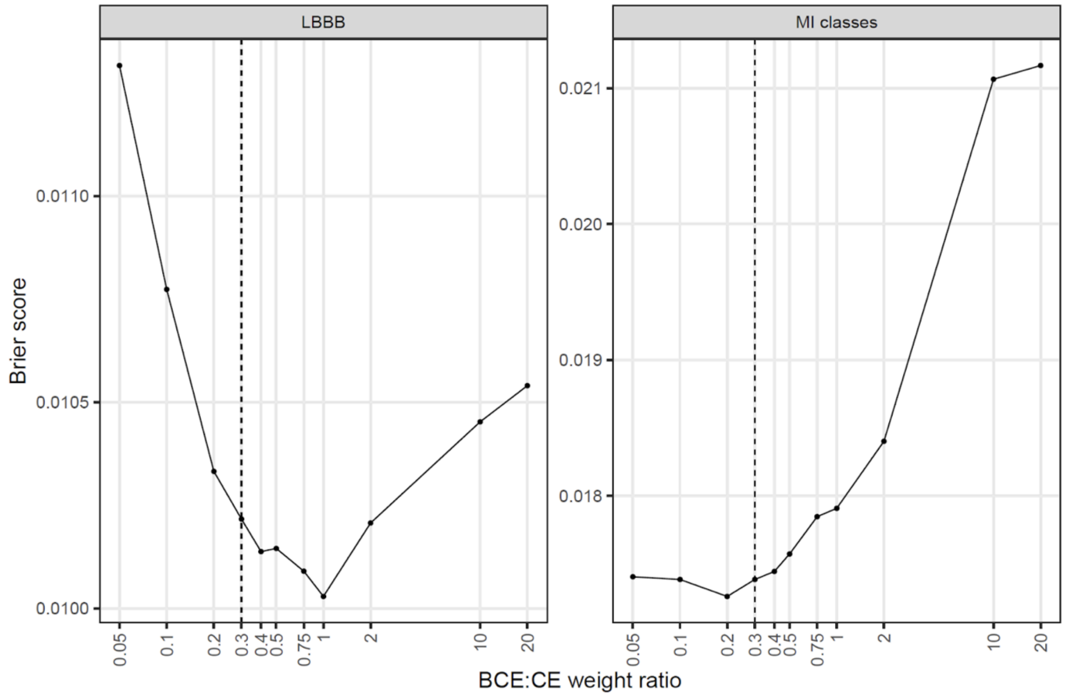

**Supplementary Figure 13.** Weighted total loss over epochs, across all five ensemble members.

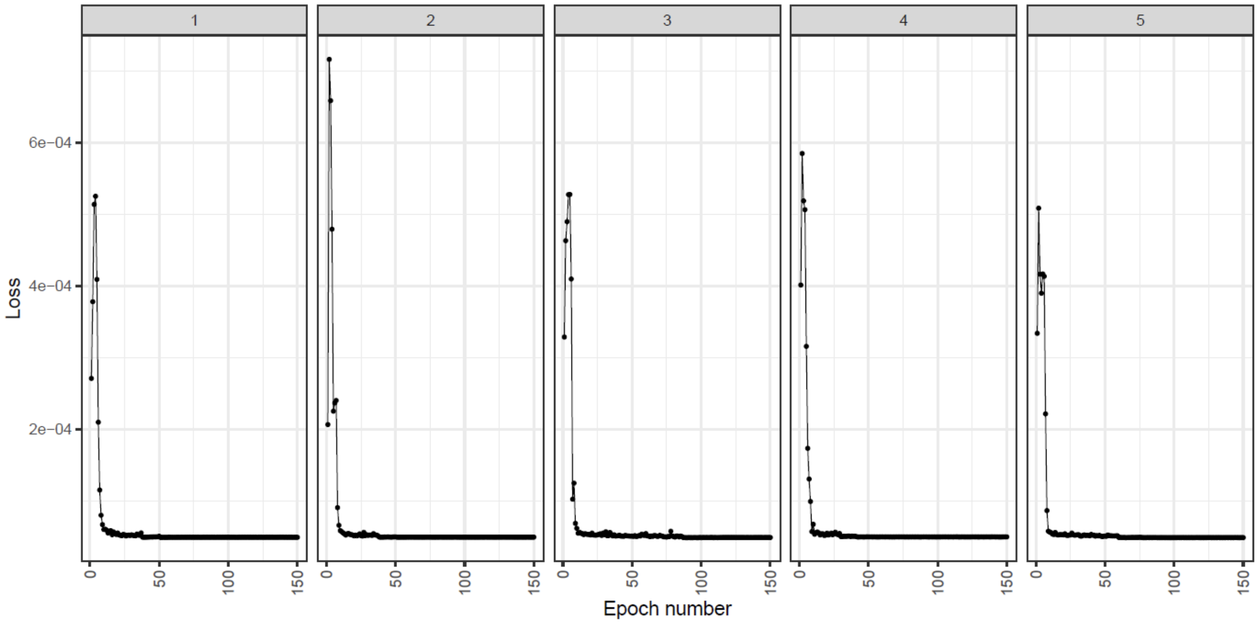
